# Multi-Center “Replica Study” Challenges the Impact of Electronic Cigarette Aerosols on Cisplatin Resistance in Head and Neck Cancer Cells

**DOI:** 10.1101/2025.01.07.25319886

**Authors:** R. Emma, G. Carota, K. Partsinevelos, S. Rust, A. Sun, A. Giordano, V. Volarevic, R. Lesmana, H. Goenawan, M. I. Barliana, A. Arsenijevic, N. Kastratovic, B. Spasic, Chiara Giardina, Miriana Cantali, R. Polosa, M. Caruso, G. Li Volti

**Affiliations:** Department of Clinical and Experimental Medicine, University of Catania, Via S. Sofia, 97, 95123, Catania (Italy); Center of Excellence for the Acceleration of Harm Reduction (CoEHAR), University of Catania, Via S. Sofia, 97, 95123, Catania (Italy); Department of Biomedical and Biotechnological Sciences, University of Catania, Via S. Sofia, 97, 95123 Catania (Italy); ECLAT Srl, spin off of the University of Catania, Via. S Sofia 89, 95123 Catania (Italy); Department of Biology, College of Science and Technology, Temple University, Philadelphia, PA 19122, USA; Sbarro Institute for Cancer Research and Molecular Medicine, Center for Biotechnology, College of Science and Technology, Temple University, Philadelphia, PA 19122, USA; Department of Medical Biotechnologies, University of Siena, Siena, Italy; Center for harm reduction of biological and chemical hazards, Faculty of Medical Sciences University of Kragujevac, 69 Svetozara Markovica 69 34000 Kragujevac, Serbia; Department of Genetics, Faculty of Medical Sciences, University of Kragujevac, 69 Svetozar Markovic Street, Kragujevac, Serbia; Department of Microbiology and Immunology, Faculty of Medical Sciences, University of Kragujevac, 69 Svetozar Markovic Street, Kragujevac, Serbia; Department of Biomedical Sciences, Faculty of Medicine, Universitas Padjadjaran, Bandung, Indonesia; Division of Biological Activity, Central Laboratory, Universitas Padjadjaran, Bandung, Indonesia; Center of Excellence for Pharmaceutical Care Innovation, Universitas Padjadjaran, Bandung, Indonesia; Department of Biological Pharmacy, Faculty of Pharmacy, Universitas Padjadjaran, Jl. Ir. Soekarno Km 21, Jatinangor, 45363, Indonesia

**Keywords:** cisplatin resistance, nicotine, electronic cigarette, chemosensitivity, head and neck squamous cell carcinoma (HNSCC)

## Abstract

**Background:** Cisplatin chemoresistance is a critical challenge in the treatment of head and neck squamous cell carcinoma (HNSCC). Since previous research has suggested that nicotine and e-cigarette (e-cig) aerosol might increase cisplatin resistance in oral cancer cells, this multicenter replication study aimed to replicate the work by Manyanga et al. (2021) and evaluate the oncologic implications of e-cigarette exposure during chemotherapy.

**Methods:** This in vitro study involved standardized and harmonized protocols in international laboratories to examine the effects of cigarette smoke (1R6F) and e-cig aerosols with different concentrations of nicotine (0, 12, and 20 mg/ml nicotine) on cisplatin sensitivity in HNSCC cell lines (SCC-25, FaDu, and UM-SCC-1). Aerosols from 1R6F smoke and e-cig vapor were collected using a smoking and vaping machine, following ISO20778:2018 and ISO20768:2018 puffing regimes. The smoke and vapor were collected in PBS and diluted to 10 puffs/5L for HNSCC cell treatment. Chemosensitivity, clonogenicity, expression of gene for repair of cisplatin-induced DNA damage and gene and protein expression of cisplatin transporters were assessed by MTS, NRU, trypan blue, PCR, and Western blot assays, respectively.

**Results:** Contrary to previous findings, exposure to e-cig aerosols did not significantly modulate cisplatin sensitivity in all cell lines. IC50 values, cytotoxicity assays, and clonogenic survival rates remained similar between e-cig treatments and cisplatin alone. Analysis of gene and protein expression revealed sporadic changes in the levels of transporters and repairs of cisplatin-induced DNA damage.

**Conclusions:** This study did not fully substantiate previous claims of increased cisplatin resistance due to e-cigarette aerosols and nicotine. The variability in gene and protein expression among different cell lines underscores the need for cautious interpretation and further investigation of the role of e-cigarette components in cancer treatment. These findings provide a critical perspective for shaping public health policies and clinical practices related to e-cigarette use during chemotherapy.

## Introduction

Cisplatin-based chemotherapy remains a cornerstone in the treatment of head and neck squamous cell carcinoma (HNSCC)(1), a prevalent malignancy characterized by poor survival rates due to late-stage diagnosis and therapy resistance. Since the coming of cisplatin as a cancer drug for head and neck cancer in the 1970s, this chemo drug has stayed key part of treatment. Now, it is linked with other kinds of treatments that make it more specific and stronger, while also boosting its healing ability and success rate (2). Yet, environmental factors and genes often lead to resistance against cisplatin. This risk harms how well the treatment works and makes the problems linked with HNSCC worse.

Using tobacco is still the top environmental risk factor for head and neck cancer, making up around 75% of all cases in a study from Western Europe (3). Smoking after getting a cancer diagnosis brings many risks, like worse response to treatment, higher chance of failure and new cancers, plus a lower quality of life. Besides where the disease is located and its stage, continued smoking is seen as the biggest negative sign for survival in people with cancer. Yet, quitting smoking has benefits that are not well recognized: many patients do not know about dangers linked to using tobacco after they have been diagnosed with cancer. Also health care workers usually do not push their patients to stop and don’t give help on quitting for those who keep using tobacco Even with the clear effect of smoking on treatment results, info about present smoking state is seldom taken in health studies (4) Many folks stop or try to stop soon after a cancer diagnosis, but still about half of those who smoke and have had cancer keep smoking. A large and strong set of studies show that keeping on with tobacco in cancer patients leads to more harm from treatments, greater chance of treatment failure, more cases of new tumors, worse life quality and shorter living time (4). Since it is hard for these people to quit smoking, and the good they might get from stopping cigarettes and other tobacco products, using another plan could make the therapy work better and improve chances for getting well. An alternative that has been available on the market for about a decade is the electronic cigarette, which gives smokers the possibility of continuing to take nicotine from a cleaner source, i.e. from an aerosol, without having to take in the by-products of tobacco combustion.

The usage of electronic cigarettes, or "e-cigarettes," which are promoted as a safer alternative to traditional smoking, has increased exponentially in recent years due to changes in nicotine use habits. However, the safety of e-cigarettes, particularly in the context of cancer treatment, remains under scrutiny. The possibility that e-cigarette aerosols could alter the effectiveness of chemotherapy, especially by enhancing cisplatin resistance in oral cancer cells, has been questioned by preliminary research, including the groundbreaking study by Manyanga et al. (2021) (5).

The study by Manyanga et al. (5) provided critical insights into how e-cigarette exposure alters the expression of cisplatin transporters, thereby increasing drug resistance. Mechanisms implicated in this resistance include changes in drug influx and efflux transporters, independent of nicotine content (6). These findings underscore the urgent need for evidence-based evaluations of e-cigarette use, particularly among cancer patients undergoing chemotherapy.

Since the spotlight was turned on the replicability crisis about a decade ago, many studies have been organized to validate the findings reported in scientific works of various nature (7). This collaborative replication study aimed to validate and expand upon the findings of Manyanga et al., investigating the effects of e-cigarette aerosols on cisplatin sensitivity in HNSCC cell lines, through the collaboration of an international team in a multicenter study (Replica Study) aimed at producing solid and credible data (8-10). By confirming or challenging these results, our work seeks to contribute to the growing body of literature assessing the oncological implications of e-cigarette use, with the ultimate goal of informing clinical practices and public health policies.

## Material and Methods

### Study design and Harmonization process

This research work represents an in vitro interlaboratory study conducted as part of the new phase of the REPLICA 2.0 project. The collaborative network for this initiative involves laboratories based in Italy (LAB-A), which serves as the coordinating center, as well as in the United States (LAB-B), Indonesia (LAB-C) and Serbia (LAB-D). In line with the guidelines for promoting transparency and openness (TOP) proposed by the Center for Open Science (https://www.cos.io/initiatives/top-guidelines), experimental protocols were harmonized among all laboratories through the development of standardized operating procedures (SOPs) for each experimental phase. To ensure consistency, all participating centers used the same cell line, exposure systems and analytical methods for endpoint assessment. LAB-A organized a harmonization meeting aimed at training the international teams and unifying the SOPs. These procedures were designed to closely mirror the original study protocols, with the exception of the aerosol generation method. In particular, non-standardized exposure conditions, which had been reported as a limitation in Manyanga et al. (5), were addressed using standardized smoking and vaping machines (LM1, LM4E/LM5E; Korber - Hamburg, Germany). Another difference from the original work is the replacement of two cell lines, WSU-HN6 (tongue) and WSU-HN6 (pharynx), as they were not commercially available. These two cell lines were replaced with SCC-25 (tongue) and FaDu (pharynx), respectively. LAB-A, LAB-B, and LAB-C conducted experiments using all the three cell lines. Whereas LAB-D conducted the experiments using only the SCC-25 cell line. We also performed the MTS assay instead of the MTT analogue and added cell viability assessment by Neutral Red Uptake (NRU) assay. Protein expression assessment by in cell western was also added, which was conducted only by LAB-C. All experiments were performed in two independent experimental sessions, with technical triplicates for each condition to ensure reproducibility and accuracy, except for the experiments conducted by LAB-A on UM-SCC-1, which were conducted in triplicate for one experimental session due to delays in cell line acquisition. The in cell western assay by LAB-C was conducted in quadruplicate for one experimental session.

### Cell culture

Following the indicated cell lines utilized by Manyanga et al. (5), human epithelial cancer cell lines from different head and neck regions were chosen for this project. SCC-25 cells (tongue; ATCC: CRL-1628) were cultured in Dulbecco’s modified Eagle’s Medium (DMEM): F-12 (ATCC: 30-2006) supplemented with 10% fetal bovine serum (FBS), 1% Penicillin/Streptomycin, and 400ng/mL hydrocortisone. FaDu cells (pharyngeal; ATCC: HTB-43) were cultured in Eagle’s Minimum Essential Medium (EMEM) (ATCC: 30-2003) supplemented with 10% FBS, and 1% Penicillin/Streptomycin. UM-SCC-1 cells (floor of mouth; Millipore: SCC070) were cultured in DMEM with 4500 mg/L glucose (Millipore: SLM-021-B) supplemented with 10% FBS, 1% Penicillin/Streptomycin, 1% Non-Essential Amino Acids (Millipore: TMS-001-C), and 1% L-Glutamine (Millipore: G7513-100ML). All the cell cultures were maintained in a humidified atmosphere at 37 °C and 5% CO_2_. The media used to prepare all the experimental conditions were supplemented with HEPES buffer (20 mM).

### Test products for smoke and aerosol generation

Smoke and aerosol were generated using a standardized experimental tobacco reference cigarette, 1R6F (Center of Tobacco Reference Products, University of Kentucky), and a commercially available e-cigarette (Joyetech eGo AIR), respectively. The eGo AIR device features a built-in 650mAh battery, an integrated 1.0-ohm coil, and a maximum power output of 12 watts, with a tank capacity of 2 ml. Aerosol generation was carried out using three commercial e-liquids with nicotine concentrations of 20, 12, and 0 mg/ml (Watson Reserve, Fashion Vape E-liquid).

### Preparation of smoke and aerosol Aqueous Extracts (AqEs)

For the 1R6F cigarette smoke exposure was used the Borgwaldt LM1 smoking machine (Borgwaldt KC GmbH). The LM1 is a linear one-port mechanical syringe-based smoking machine that delivers undiluted smoke. The 1R6F reference cigarettes were conditioned before use for at least 48 hours at 22 ± 1 °C and 60 ± 3% relative humidity, in accordance with ISO 3402:2023 (11). Subsequently, 1R6F were smoked following the Health Canada Intense (HCI) regimen (55 ml puff volume, 2 seconds puff duration, 30 seconds puff frequency, with bell shaped profile, 27.5 ml/s puff velocity, and the hole vents blocked), accredited under ISO 20778:2018 (12). Laboratory conditions were monitored using temperature and humidity sensors before and during the exposure, ensuring a test atmosphere with 60% (± 5%) relative humidity and a temperature of 22 °C (± 2 °C), also following ISO 3402:2023 (11).

E-cigs were used according to the manufacturer’s instructions. Before each exposure, they were fully charged, cleaned, and loaded with fresh consumables. For the aerosol exposure was used the Borgwaldt LM4E/LM5E vaping machine (Korber GmbH). The three different e-liquids loaded into the e-cigs were vaped according to the “CORESTA Reference Method n. 81” (CRM81) regimen (55 ml puff volume, 3 seconds puff duration, 30 seconds puff frequency, with square shaped profile, and 18.3 ml/s puff velocity), accredited into ISO 20768:2018 (13). During the exposure, a relative humidity of 40–70% and a temperature of 15–25 °C (±2 °C) were maintained in the laboratory, also in accordance with ISO 20768:2018.

Aqueous extracts (AqEs) were prepared by bubbling 9 puffs of smoke or aerosol into 45 ml of phosphate-buffered saline (PBS) contained in an impinger, resulting in a concentration of 0.2 puffs/ml. The stock AqEs were diluted 1:100 in order to achieve a final concentration of 0.002 puffs/ml (equivalent to 10 puffs/5L), which corresponds to the concentration used in the Manyanga’s study (5). Nicotine was quantified in each stock AqE by reversed-phase HPLC as reported in the supplementary materials, and results were reported in table S1.

### Chemosensitivity assay by MTS and NRU

MTS and the Neutral Red Uptake (NRU) assays were used for determining the cell viability after treatment with Aqueous Extract (AqE) of 1R6F smoke and e-cigs aerosol, in presence and absence of Cisplatin at different concentrations. Cells were seeded 24h before treatment in 96-well plates at a density of 5 × 10^3^ (SCC-25), 3 x 10^3^ (FaDu), and 1.25 x 10^3^ (UM-SCC-1) cells per well. The next day, the cells were treated with AqEs (from 1R6F smoke and e-cigs aerosol with 20, 12 and 0 mg/ml of Nicotine) at a concentration of 0.002 puffs/ml. After 48h, the treatment was replaced with new AqEs (0.002 puffs/ml) in combination with different concentrations of Cisplatin (100, 10, 1, 0.1, 0.01 μM). After 48h, the MTS and NRU assays were performed following the manufacturer’s instructions. After every treatment, a microplate sealing membrane (Diversified Biotech Breath-EASIER BERM-2000) was applied to the plate.

#### MTS assay

CellTiter 96® AQueous One Solution Cell Proliferation Assay (Promega: G3580) was used by LAB-A, LAB-B, and LAB-D. In brief, MTS assay was performed by replacing all the treatments with fresh medium, and by adding 20 μL of MTS Solution to each well. The plates were incubated at 37 °C for 3 hours in a humidified atmosphere of 5% CO_2_. A microplate reader was used to detect the optical density of the MTS solution (490 nm filter). CellTiter-Glo® Luminescent Cell Viability Assay (Promega: G7572) was used by LAB-C. A volume of CellTiter-Glo® Reagent equal to the volume of the treatment present in each well was added. After 2 minutes on an orbital shaker, the plates were incubated at room temperature for 10 minutes. The generated luminescent signal was recorded using a luminometer microplate reader.

#### NRU assay

The NRU vital dye staining by Sigma Aldrich (#N2889) was used by all the laboratory. The day before NRU assay, the NR solution was prepared in medium at ratio 1:65 (0.05 g/L) plus HEPES buffer at 20 mM and placed in incubator at 37 °C 5% CO_2_. The day of NRU assay, the NR solution was filtered prior to use (0.2 μm filter). After treatment removal, the cells were washed with PBS. Then, 150 μL of the filtered NR solution were added to each well. The plates were incubated at 37°C for 3 hours in a humidified atmosphere of 5% CO_2_. After incubation, NRU solution was removed, and the cells were washed with PBS. The NR Destain Solution was prepared immediately before the use (Ratio: [ethanol : glacial acetic acid : distilled water] [50 : 1 : 49]) and added in each well. After 10 minutes of incubation on a plate shaker, the optical density was detected by a microplate reader with a filter of 540 nm and a background filter of 630 nm.

### Trypan blue exclusion assay

Cell viability after treatment with 1R6F or e-cig AqEs, both with and without Cisplatin, was also assessed using the Trypan blue exclusion assay. Cells were seeded 24h before treatment in 48-well plates at a density of 10 × 10^3^ (SCC-25), 6 x 10^3^ (FaDu), and 20 x 10^3^ (UM-SCC-1) cells per well. The day after, the cells were treated with 1R6F or e-cig (20, 12 and 0 mg/ml of Nicotine) AqEs at a concentration of 0.002 puffs/ml. After 48h, the treatment was replaced with fresh AqEs (0.002 puffs/mL), with or without 2.5 µM Cisplatin. After another 48h, floating cells were collected, and adherent cells were harvested using trypsin. The two cell populations were then mixed, pelleted, stained with 0.4% Trypan blue (Gibco) at a 1:1 ratio, and counted using a Neubauer chamber.

### Clonogenic survival assay

Cell seeding and treatment procedures were carried out as previously described for the Trypan blue exclusion assay. Afterwards, to assess colony survival, adherent cells were trypsinized, pelleted, counted, and seeded into 6-well plates at a density of 10^3^ (SCC-25, FaDu, and UM-SCC-1) cells per well. Cells were cultured for 7 – 15 days to allow colony formation, with the media changed every 2-3 days. When colony growth was sufficient, methanol was added in each well for 15 minutes in order to fix the cells. Subsequently, the cells were stained with 0.5% Crystal Violet (Sigma) in 25% methanol – 75% distilled H_2_O for 10 minutes. The colonies were then washed 4 times with distilled H_2_O and air-dried. Colonies formed by 50 or more cells were counted under a microscope. Plating efficacy (PE) and the surviving fraction (SF) values were calculated as follow (14):

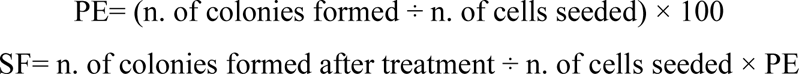

### Protein expression by Western Blot and In-cell Western

The protein expression of Cisplatin transporters after cells treatment with AqEs (PBS bubbled with 1R6F smoke and e-cigs aerosol with 20, 12 and 0 mg/ml of Nicotine) was assessed by Western Blot analysis (LAB-A, LAB-B, and LAB-D) and by In-cell Western (LAB-C).

#### Western Blot

The day prior to the treatment, the cells were seeded in T-25 flasks at a density of 2 × 10^5^ (SCC-25), 1 x 10^6^ (FaDu), and 3 x 10^5^ (UM-SCC-1) cells per flask. After 24h, the cells were treated with the different AqEs at a concentration of 0.002 puffs/ml. 48 hours after exposure to AqEs, cells were trypsinized, pelleted, washed with ice-cold PBS, and then resuspended in radioimmunoprecipitation assay (RIPA) buffer (Sigma-Aldrich: R0278-50ML) supplemented with 1X Protein Inhibitor Cocktail (Sigma-Aldrich). The protein quantification was performed with the Protein Assay Dye Reagent Concentrate (Biorad) using bovine serum albumin (BSA) as standard. The total protein content was quantified and an amount of 30-50 μg for each sample was denatured for 10 min at 70°C in an appropriate loading buffer composed by NuPAGE™ LDS Sample Buffer (Invitrogen) and Bolt™ Sample Reducing Agent (Invitrogen). The separation of proteins was performed by electrophoresis using a 4-12% polyacrylamide gel (Bolt™ Bis-Tris Plus Mini Protein Gels, 4-12%, 1.0 mm, WedgeWell™ format) followed by the electro-transfer of proteins onto the nitrocellulose membrane (Power Blotter Select Transfer Stacks). Subsequently, the membranes were blocked with 5% BSA solution for 1h at room temperature. After blocking the nonspecific sites, the membranes were incubated overnight at room temperature with the following primary antibodies resuspended in 5% BSA solution: anti-ATP7A (1:100; Santa Cruz Biotechnologies, Texas, USA, sc-376467), anti-CTR1 (1:40.000; Abcam, Massachusetts, USA, #ab129067), anti-ABCG2 (1:100; Cell Signaling Technology, Massachusetts, USA, #42078), and β-actin as a loading control (1:20.000; Cell Signaling Technology, Massachusetts, USA, #8457). After incubation, the membranes were washed three times with Tris-buffered saline with 0.1% Tween 20 (TBS-T), and subsequently incubated for 1h at room temperature with the following secondary antibodies resuspended in 5% BSA solution: rabbit anti-mouse IgG (1:10.000; Abcam, Massachusetts, USA, #ab6728) and goat anti-Rabbit IgG (1:10.000; Cell Signaling Technology, Massachusetts, USA, #7074). The membranes were then washed four times with TBS-T. Finally, the protein bands were visualized using the Clarity Max Western ECL Substrate (Biorad: 1705062) and detected by Li-Cor C-DiGit® Chemiluminescent Western Blot Scanner. The protein levels were quantified by densitometric analysis using the ImageJ software, while data were normalized on β-actin expression levels.

#### In-cell Western

SCC-25, FaDu, and UM-SCC-1 cells were seeded at a density of 5×10^3^, 3×10^3^, and 1.25×10^3^ cells/well, respectively, in a 96-well black plate (Thermo Scientific, USA) and incubated at 37 °C in a 5% CO_2_. After 24h of incubation, the cells were treated with the different conditions of AqE (1R6F smoke and e-cig aerosol with 20, 12, and 0 mg/ml of Nicotine) at a concentration of 0.002 puff/ml, for 24h. After incubation, the cells were fixed with 3,7% formaldehyde (150 μl) (Merck Cat No. 1040031000, Germany) for 20 minutes at room temperature. The cells were then permeabilized using 0.1% Triton X-100 in TBS (150 μl) for 20 minutes, by gently agitating on a microplate shaker. Then, the cells were blocked with the Intercept Blocking Buffer (LICORbio, P/N: 927-70001) for 30 minutes. After blocking buffer removal, the cells were incubated with 50 μl of the following primary antibodies: CTR1 (#13086), ATP7A (Santa Cruz Biotechnology), ABCG2 (D5V2K Rabbit mAb #42078), and ß-actin (D6A8 Rabbit mAb #8457). The antibodies were prepared in PBS-Tween with 0,1 mg of BSA at 1:1000 dilution. The plate was incubated overnight at 4 °C in the dark. After incubation, the primary antibodies were removed from the wells. All the wells were washed four times with washing solution (150 μl) at room temperature by gently agitating with microplate shaker. Subsequently, the plate was dried and incubated with 50 μl of secondary antibodies: IRDye® 800 CW Goat anti-Rabbit IgG Secondary Antibody 925-32211, IRDye® 800 CW Goat anti-Mouse IgG Secondary Antibody 925-32210, and IRDye® 680 RD Goat anti-Rabbit IgG Secondary Antibody 925-68071. The plate was incubated in the dark for 1h at room temperature. The secondary antibodies were removed, and the wells were washed four times. The protein expression was evaluated with Odyssey® CLx Imaging System (Licor Biosciences, US) with 169 μm resolutions, Lowest Quality, 4.0 mm Focus offset, and 800 wavelenght Intensity. For ß-actin detection, 680 wavelenght Intensity was used. Untreated cell monolayers were used for subtracting unspecific and/or background fluorescent signals. All the data were acquired by Licor Image Studio Software (version 3.1), while recorded values were then exported into Excel (Microsoft, US, version 2010).

### Gene expression by Real-Time PCR

Real-Time PCR was employed to evaluate the gene expression of membrane transport proteins involved in Cisplatin uptake and efflux in cells exposed to cigarette smoke and e-cig aerosol (Table S2). The day prior to the treatment with AqEs, cells were seeded in 12-well plates at a density of 50 x 10^3^ (SCC-25), 30 x 10^3^ (FaDu), and 40 x 10^3^ (UM-SCC-1) cells/well. After 24h, cells were treated with AqE from 1R6F smoke and e-cigs aerosol with 20, 12 and 0 mg/ml of Nicotine, at a concentration of 0.002 puff/ml, and then incubated for 48h. After incubation, cells were detached and collected by using the RLT lysis buffer (Qiagen) supplemented with β-mercaptoethanol, and the RNA was then extracted by RNeasy Mini Kit (Qiagen), following the manufacturer’s instructions. RNA content and related purity were evaluated by spectrophotometric absorbance (A260/A280 nm ratio). The cDNA synthesis via reverse transcription from the isolated RNA was carried out with the QuantiTect Reverse Transcription Kit (Qiagen) according to the manufacturer’s protocol. Quantitative Real-Time PCR was performed using the KAPA SYBR FAST Universal (KAPA Biosystems), while the obtained mRNA expression levels were normalized with β-actin as housekeeping gene, by using a comparative 2^−ΔΔ*C*t^ method.

### Statistics

All raw data were organized and processed using Microsoft Excel. Data were analyzed following predefined criteria to identify outliers or anomalous results. Some experiments were excluded from the analysis under the following parameters: if an entire condition showed an anomalous result compared with the other experimental replicates and if the technical cause was not identified. Outlier detection was performed using the ROUT (robust regression-based outlier rejection) test. To assess the normality or skewness of the data distribution, the Shapiro-Wilk test was applied. Symmetrical data were expressed as mean ± standard error of the mean (SEM), while skewed data were represented as median (95% confidence interval - 95% CI). The intraclass correlation coefficient (ICC) was calculated using a two-way mixed-effects model with absolute agreement to assess the consistency and repeatability of intrasession measurements across laboratories. ICC calculations were performed with R software, version 4.2.3 (2023-03-15). To identify statistically significant differences between study groups, ANOVA or Kruskal-Wallis test was used. Further comparisons between groups were conducted using post-hoc tests, including Tukey’s test or Dunn’s test. IC50 values for the tested products were determined by fitting a sigmoidal dose-response curve with variable slope, using nonlinear regression to identify the best parameters and compare the IC50 values. Statistical significance was set at p < 0.05 for all analyses. GraphPad Prism 8 software was used for data analysis and generation of graphs unless otherwise stated.

## Results

### Reproducibility across all laboratories

To assess the reproducibility of our results, we calculated the intraclass correlation coefficient (ICC) for cell viability, cell death, chemosensitivity, clonogenic, gene expression and western blot results from the different laboratories. The ICC values, together with their 95% confidence intervals and corresponding p values, are given in the supplementary materials (Tables S3-S8).

#### Cell viability data (MTS and NRU)

The reproducibility of cytotoxicity assays was evaluated in multiple laboratories using MTS and NRU assays. ICC values indicated moderate to high agreement depending on cell line and treatment (Table S1). For MTS data, reproducibility was highest for SCC-25 with 1R6F treatment (ICC = 0.884, p < 0.0001), but decreased significantly for e-cig treatments (ICC = 0.556, p < 0.0001). Similar trends were observed for the other cell lines, with moderate ICCs for FaDu (0.609 for 1R6F; 0.506 for e-cig) and UM-SCC-1 (0.598 for 1R6F; 0.574 for e-cig). For NRU data, higher ICC values were observed for FaDu in both treatments (ICC = 0.932 for 1R6F; 0.906 for e-cig, both p < 0.0001), while SCC-25 and UM-SCC-1 showed moderate reproducibility, with ICC values ranging from 0.500 to 0.665.

These results highlight that the NRU assay generally provided better interlaboratory reproducibility than the MTS assay, particularly for FaDu cells.

#### Trypan Blue data

Trypan Blue assay results revealed varying levels of reproducibility, with the lowest ICCs observed for the SCC-25 cell line (Table S2). For this cell line data, the ICC values were 0.154 (1R6F) and 0.119 (e-cig), reflecting poor if significant agreement (p = 0.0022 and p = 0.0005, respectively). For the FaDu cell line, reproducibility improved significantly, with ICC of 0.534 (1R6F) and 0.648 (e-cig) (both p < 0.0001). Data from UM-SCC-1 showed moderate to high agreement, with ICC values of 0.634 (1R6F) and 0.826 (e-cig), both statistically significant (p = 0.0005 and p < 0.0001, respectively).

The Trypan Blue assay showed weaker reproducibility for SCC-25 than FaDu and UM-SCC-1, suggesting that this assay is more sensitive to experimental conditions. While the Trypan Blue exclusion assay is a reliable method for validating results from MTS or NRU assays in small-scale experiments, its manual implementation at a larger scale becomes labor-intensive and time-consuming. This not only complicates the process but also makes it challenging to complete the assay within a defined timeframe, potentially compromising the results.

#### Chemosensitivity data

Chemosensitivity assays using the MTS and NRU assays showed excellent inter-laboratory reproducibility (Table S3). For MTS data, SCC-25 revealed an ICC of 0.943 (1R6F) and 0.895 (e-cig), both indicating near perfect agreement (p < 0.0001). Similar trends were observed for FaDu (ICC = 0.899 for 1R6F; 0.882 for e-cig) and UM-SCC-1 (ICC = 0.867 for 1R6F; 0.796 for e-cig). For NRU data, even higher ICC values were observed, particularly for FaDu (ICC = 0.942 for 1R6F; 0.949 for e-cig) and UM-SCC-1 (ICC = 0.885 for 1R6F; 0.894 for e-cig).

The high ICCs suggest strong reproducibility of chemosensitivity assays, especially with NRU protocols, among all cell lines.

#### Clonogenic data

The clonogenic assay showed variable reproducibility depending on treatment and cell line (Table S4). SCC-25 showed poor concordance, with ICC of 0.154 (1R6F) and 0.119 (e-cig) although this was significant (p = 0.0022 and p = 0.0005, respectively). Data from FaDu presented moderate reproducibility for both 1R6F (ICC = 0.534, p < 0.0001) and e-cig aerosols (ICC = 0.648, p < 0.0001). UM-SCC-1 showed better reproducibility, with ICC of 0.634 (1R6F) and 0.826 (e-cig) (both p < 0.0001).

These results indicate that the clonogenic assay was more reproducible for FaDu and UM-SCC-1, while the reproducibility of SCC-25 remains limited.

#### Gene expression data

The reproducibility of gene expression measurements was evaluated for major cisplatin transporter genes, including XPA, MMS19, ERCC1, ATP7A. Reproducibility was not evaluated for the other genes because their expression was conducted only by LAB-A. ICC values for gene expression data varied widely (Table S5). For SCC-25, ICC was high for MMS19 (ICC = 0.360, p < 0.0001) and moderate for ERCC1 (ICC = 0.171, p = 0.0187), but very low for XPA (ICC = 0.0163, p = 0.357). FaDu showed poor agreement for MMS19 and XPA (negative ICC values), while the reproducibility of ERCC1 was moderate (ICC = 0.283, p = 0.0162). In UM-SCC-1, ICC values were low to moderate, with the highest agreement observed for ERCC1 (ICC = 0.151, p = 0.0511).

Gene expression assays showed inconsistent reproducibility, particularly for FaDu and UM-SCC-1, highlighting the very high sensitivity of this assay to experimental variables.

#### Western blot data

The reproducibility of protein expression data from Western blot analysis was generally low (Table S6). For SCC-25 data, ICC values were poor for ATP7A (ICC = 0.132, p = 0.0751) and CTR1 (ICC = 0.120, p = 0.159). Negative ICC values were observed for ABCG2, reflecting substantial interlaboratory variability. Western blot analysis demonstrated poor overall reproducibility, supporting the reputation of this assay as a difficult and nonreproducible technique.

### Evaluation of cell viability and death induced by cisplatin in combination with 1R6F and e-cigarette aerosol

To evaluate cytotoxic effects and cell viability in response to cisplatin treatment (10 μM for MTS and NRU and 2.5 μ for trypan blue) and its combination with cigarette smoke extract (1R6F) and e-cigarette (e-cig) aerosol at different nicotine concentrations, we conducted MTS, Neutral Red Uptake (NRU) and Trypan Blue exclusion assays on three oral cancer cell lines: SCC-25 (Fig. 1), FaDu (Fig. 2) and UM-SCC1 (Fig. 3). Overall, our findings across all three oral cancer cell lines (SCC-25, FaDu, and UM-SCC-1) indicate that the addition of 1R6F cigarette smoke extract or e-cig aerosols at varying nicotine concentrations did not impact the cytotoxicity or cell death induced by cisplatin treatment. Unlike the results reported by Manyanga et al. (5), we observed no significant modulation of cisplatin sensitivity in response to these exposures. Although some conditions appear to show variability with respect to cisplatin, as with trypan blue in UM-SCC-1 cells, these variations are not statistically significant.

**Figure 1.**
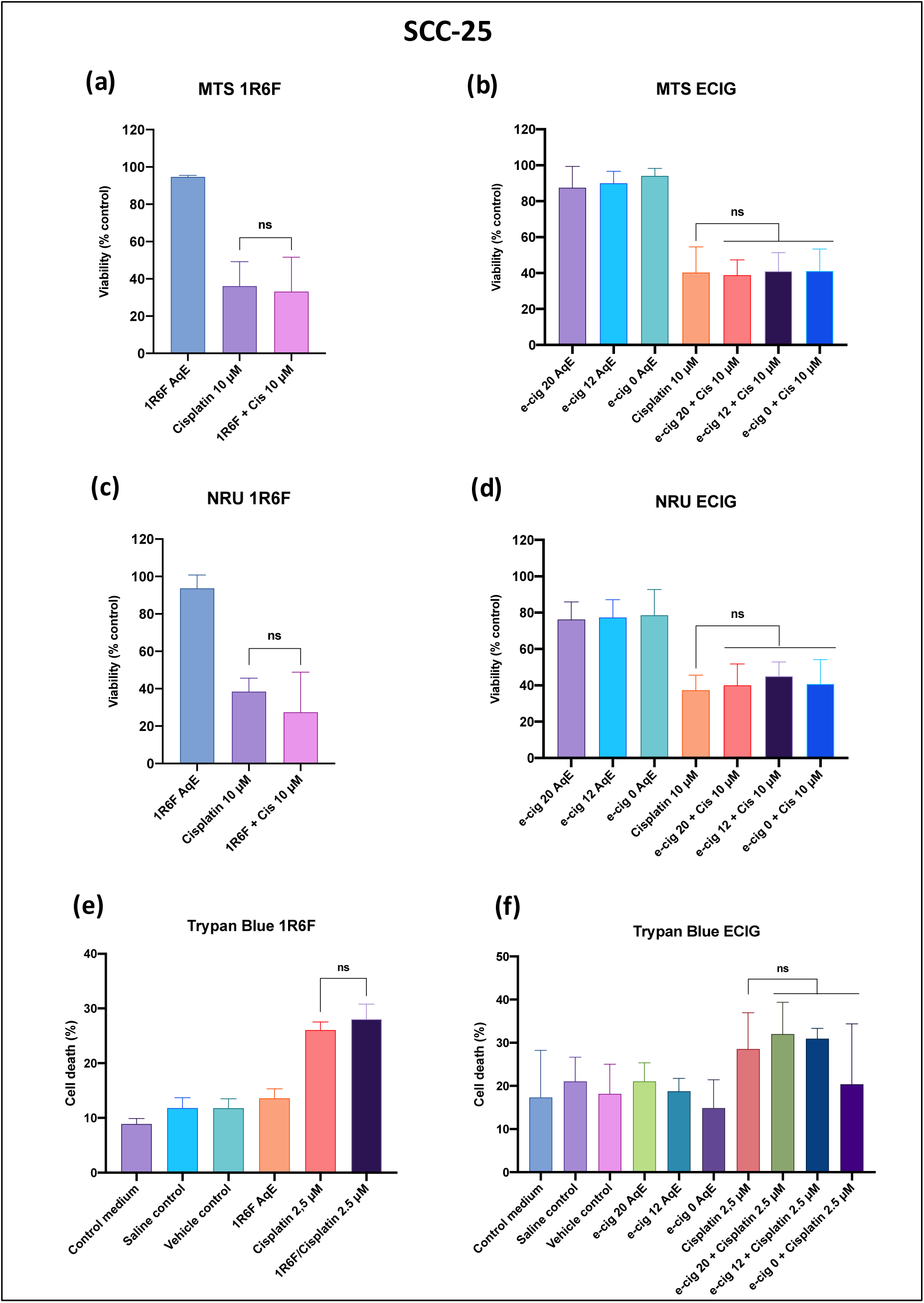
Evaluation of cell viability (MTS and NRU) and cell mortality (Trypan Blue exclusion assay) in SCC-25 cell line in response to cisplatin treatment and its combination with cigarette smoke extract (1R6F) and electronic cigarette (e-cig) aerosols. Data in the panel (a), (b), (c), (d), and (f) were presented as median and CI 95% and were analyzed by using Kruskal-Wallis test followed by Dunn’s test for multiple comparisons. Data in the panel (e) were presented as mean and SEM, and were analyzed by using ANOVA followed by Tukey’s test for multiple comparisons Ns: not significant.

**Figure 2.**
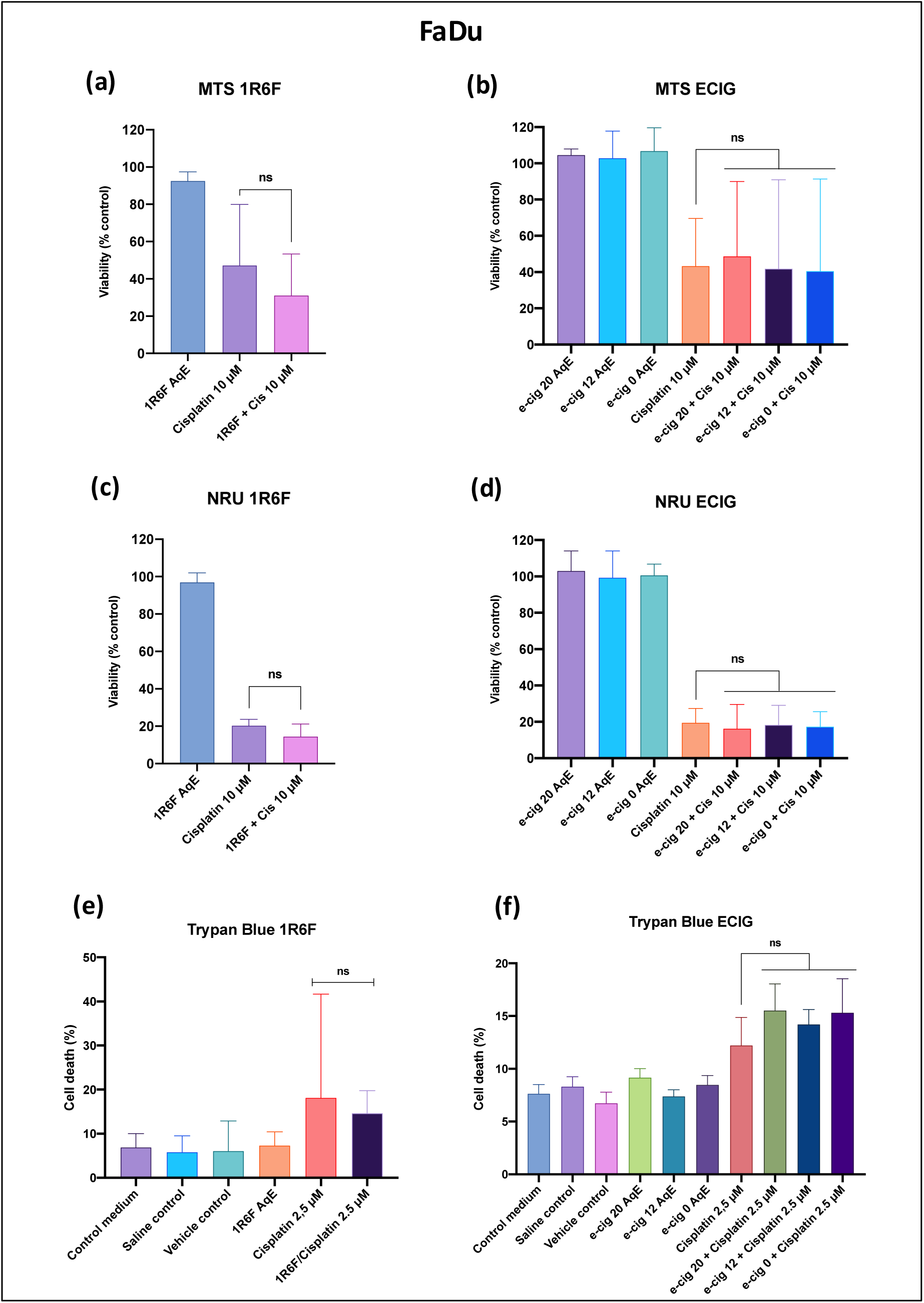
Evaluation of cell viability (MTS and NRU) and cell mortality (Trypan Blue exclusion assay) in FaDu cell line in response to cisplatin treatment and its combination with cigarette smoke extract (1R6F) and electronic cigarette (e-cig) aerosols. Data in the panel (a), (b), (c), (d), and (e) were presented as median and CI 95% and were analyzed by using Kruskal-Wallis test followed by Dunn’s test for multiple comparisons. Data in the panel (f) were presented as mean and SEM, and were analyzed by using ANOVA followed by Tukey’s test for multiple comparisons Ns: not significant.

**Figure 3.**
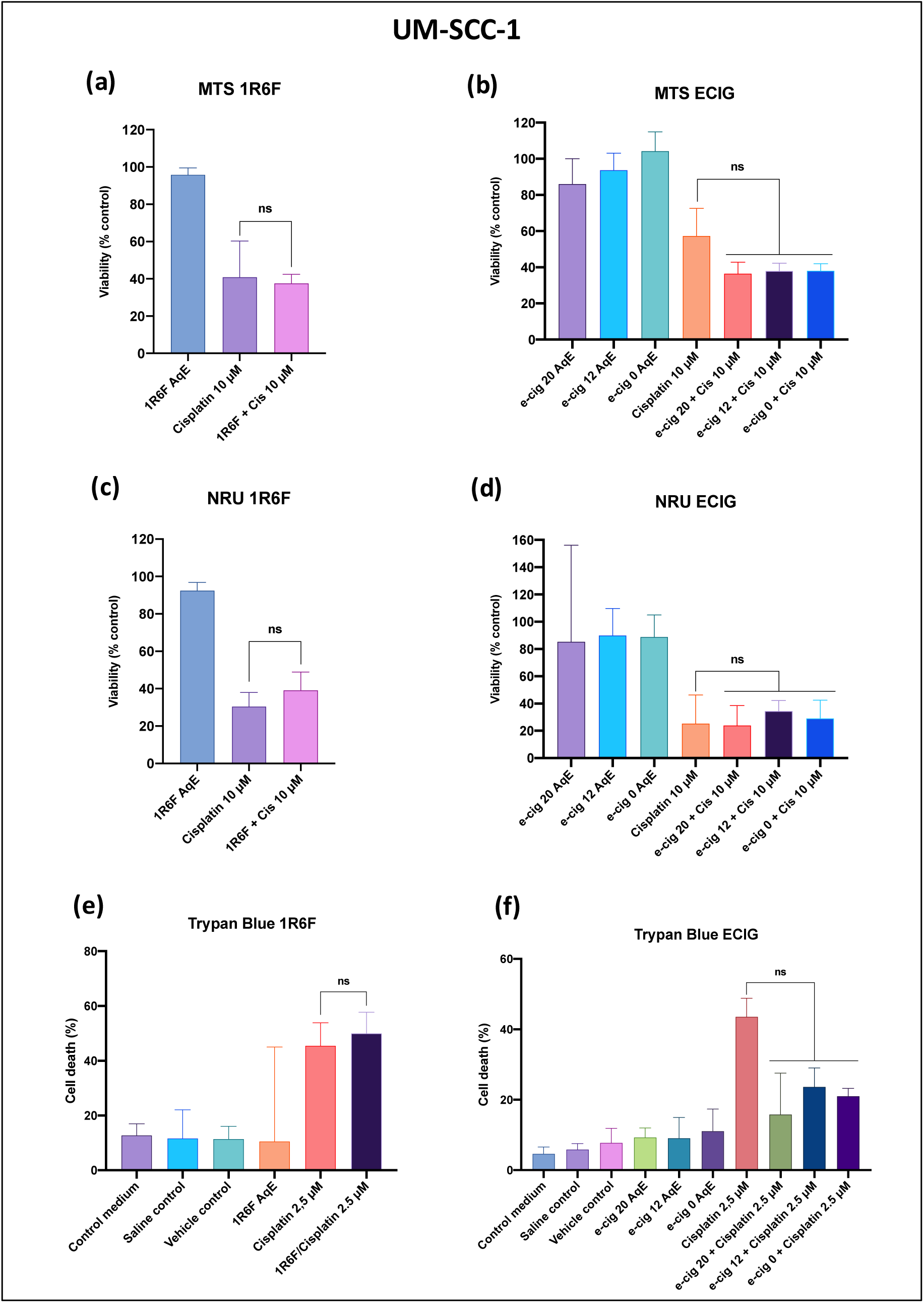
Evaluation of cell viability (MTS and NRU) and cell mortality (Trypan Blue exclusion assay) in FaDu cell line in response to cisplatin treatment and its combination with cigarette smoke extract (1R6F) and electronic cigarette (e-cig) aerosols. Data in the panel (a), (b), (d), (e) and (f) were presented as median and CI 95% and were analyzed by using Kruskal-Wallis test followed by Dunn’s test for multiple comparisons. Data in the panel (c) were presented as mean and SEM, and were analyzed by using ANOVA followed by Tukey’s test for multiple comparisons. ns: not significant.

### Evaluation of cisplatin required to induce a 50% reduction in cell growth (IC50) in combination with 1R6F and e-cigarette aerosol

The chemosensitivity of SCC-25, FaDu, and UM-SCC-1 cell lines to cisplatin was evaluated using the MTS and NRU assays. The MTS assay results revealed significant differences in drug efficacy (Fig. 4). For the SCC-25 cell line, the IC50 values for cisplatin and 1R6F were 8.09 mM and 6.39 mM, respectively, with a p-value of 0.0589, indicating an increasing trend in sensitivity with 1R6F. For SCC-25 exposed to e-cig, the IC50 values for cisplatin, e-cig 0 nic, e-cig 12 nic, and e-cig 20 nic were 10.63 mM, 10.94 mM, 10.02 mM, and 8.77 mM, respectively, with a p-value of 0.359, suggesting no significant difference between treatments. The FaDu cell line showed marked sensitivity to 1R6F (IC50= 5.81 mM) compared with cisplatin (IC50= 10.68 µM), with a highly significant p-value of 0.0007. In contrast, the FaDu exposed to e-cig showed IC50 values of 10.99 µM for cisplatin and 7.40 mM, 9.28 mM, and 8.75 mM for e-cig 0 nic, e-cig 12 nic, and e-cig 20 nic, respectively, with a p-value of 0.183, indicating no significant differences. For the UM-SCC-1 cells, no significant difference was observed between the IC50 of cisplatin and the IC50 of 1R6F cotreatment. Instead, the UM-SCC-1 exposed to e-cig demonstrated significant changes in sensitivity, with IC50 values of 9.16 mM for cisplatin and 7.96 mM, 8.56 mM, and 8.22 mM for e-cig 0 nic, e-cig 12 nic, and e-cig 20 nic, respectively, with a p-value < 0.0001.

**Figure 4.**
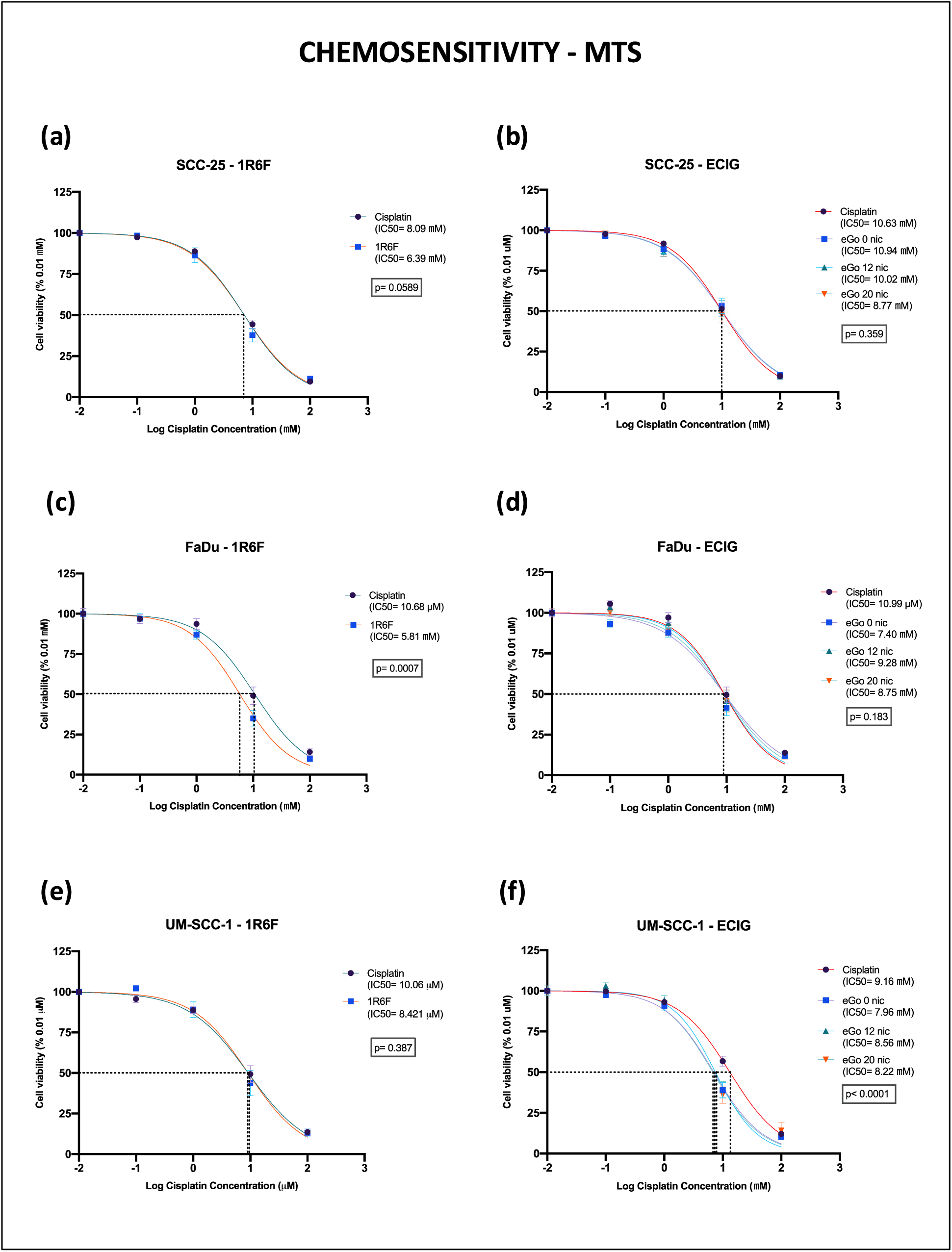
Chemosensitivity of SCC-25, FaDu, and UM-SCC-1 cell lines to cisplatin and cotreatment with 1R6F and e-cigarette aerosols evaluated using the MTS assay.

Similar results were observed when chemosensitivity was assessed by the NRU assay (Fig. 5). For the SCC-25 cell line, no significant difference was observed between the IC50 values of cisplatin and 1R6F, 6.91 mM and 5.74 mM, respectively (p = 0.259). In the evaluation of SCC-25 exposed to e-cig, the IC50 values for cisplatin, e-cig 0 nic, e-cig 12 nic, and e-cig 20 nic were 8.79 mM, 10.6 mM, 10.39 mM, and 8.96 mM, respectively, with a p-value of 0.586, suggesting no significant difference between treatments. The FaDu cell line showed significant sensitivity to 1R6F (IC50 = 2.80 mM) compared with cisplatin (IC50 = 3.90 mM), with a p-value of 0.015. In the FaDu e-cig group, the IC50 values were 3.89 mM for cisplatin and 2.88 mM, 4.07 mM, and 3.55 mM for e-cig 0 nic, e-cig 12 nic, and e-cig 20 nic, respectively, with a p-value of 0.152, indicating no significant differences. The UM-SCC-1 cell line showed IC50 values of 6.09 mM for cisplatin and 5.42 mM for 1R6F, with a p-value of 0.665, indicating no significant difference. In the UM-SCC-1 exposed to e-cig, the IC50 values were 5.10 mM for cisplatin and 6.15 mM, 4.60 mM, and 3.55 mM for e-cig 0 nic, e-cig 12 nic, and e-cig 20 nic, respectively, with a p-value of 0.123.

**Figure 5.**
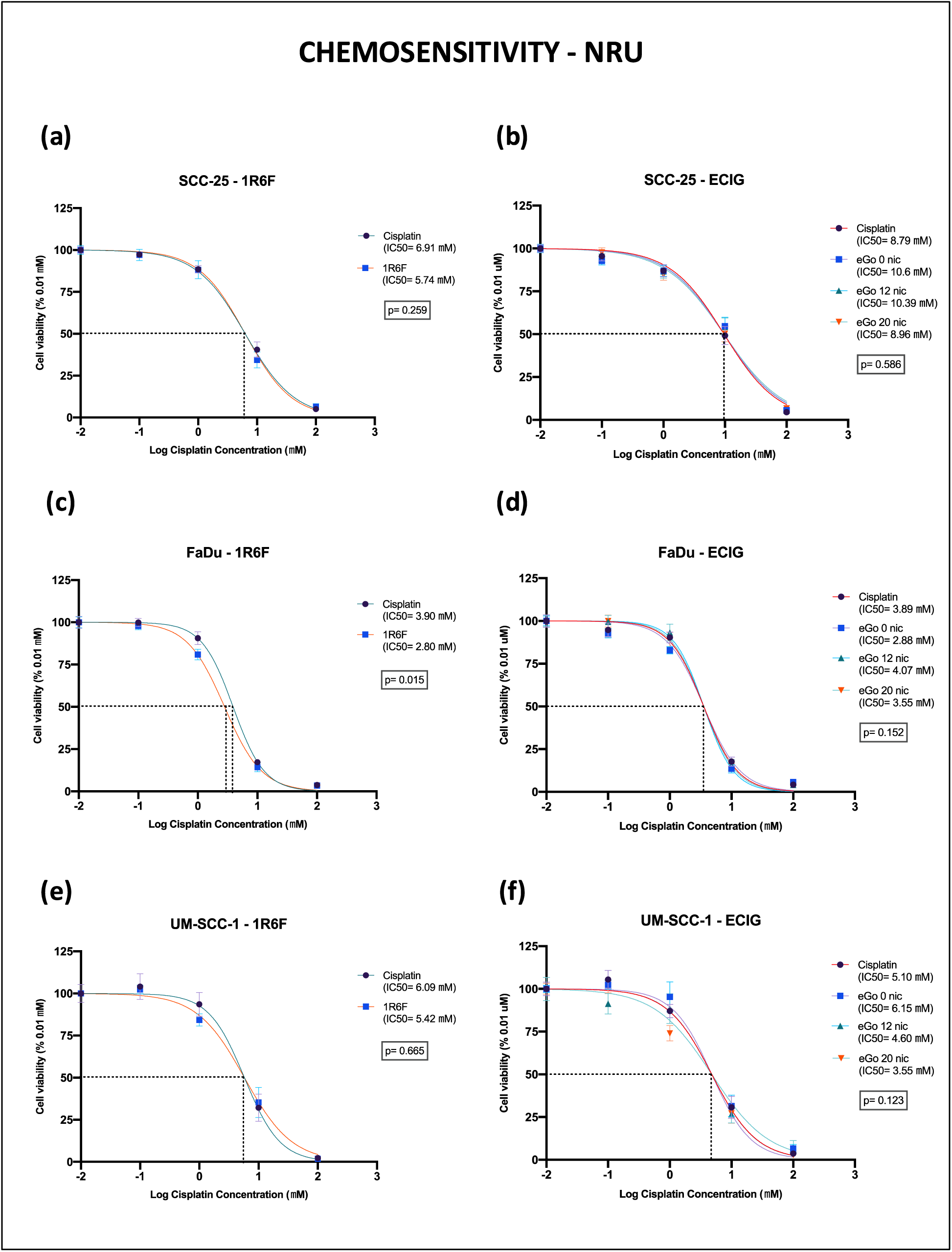
Chemosensitivity of SCC-25, FaDu, and UM-SCC-1 cell lines to cisplatin and cotreatment with 1R6F and e-cigarette aerosols evaluated using the NRU assay.

### Clonogenic survival after cisplatin treatment in combination with 1R6F and e-cigarette aerosol

The results of the clonogenic assay are shown in Figure 6 for the three different cell lines: SCC-25 (Fig. 6 a and b), FaDu (Fig. 6 c and d) and UM-SCC-1 (Fig. 6 e and f). In contrast to the work of Manyanga et al (5), our data indicate no interaction between cisplatin alone and cisplatin combined with 1R6F and e-cigarette extracts in the formation of colonies. No or few colonies were observed with cisplatin treatment alone and in combination with 1R6F and e-cig extracts. Instead, the exposure to 1R6F and e-cig extracts without the cisplatin cotreatment did not affect the reproductive viability in terms of clonogenic survival for all the cancer cell lines.

**Figure 6.**
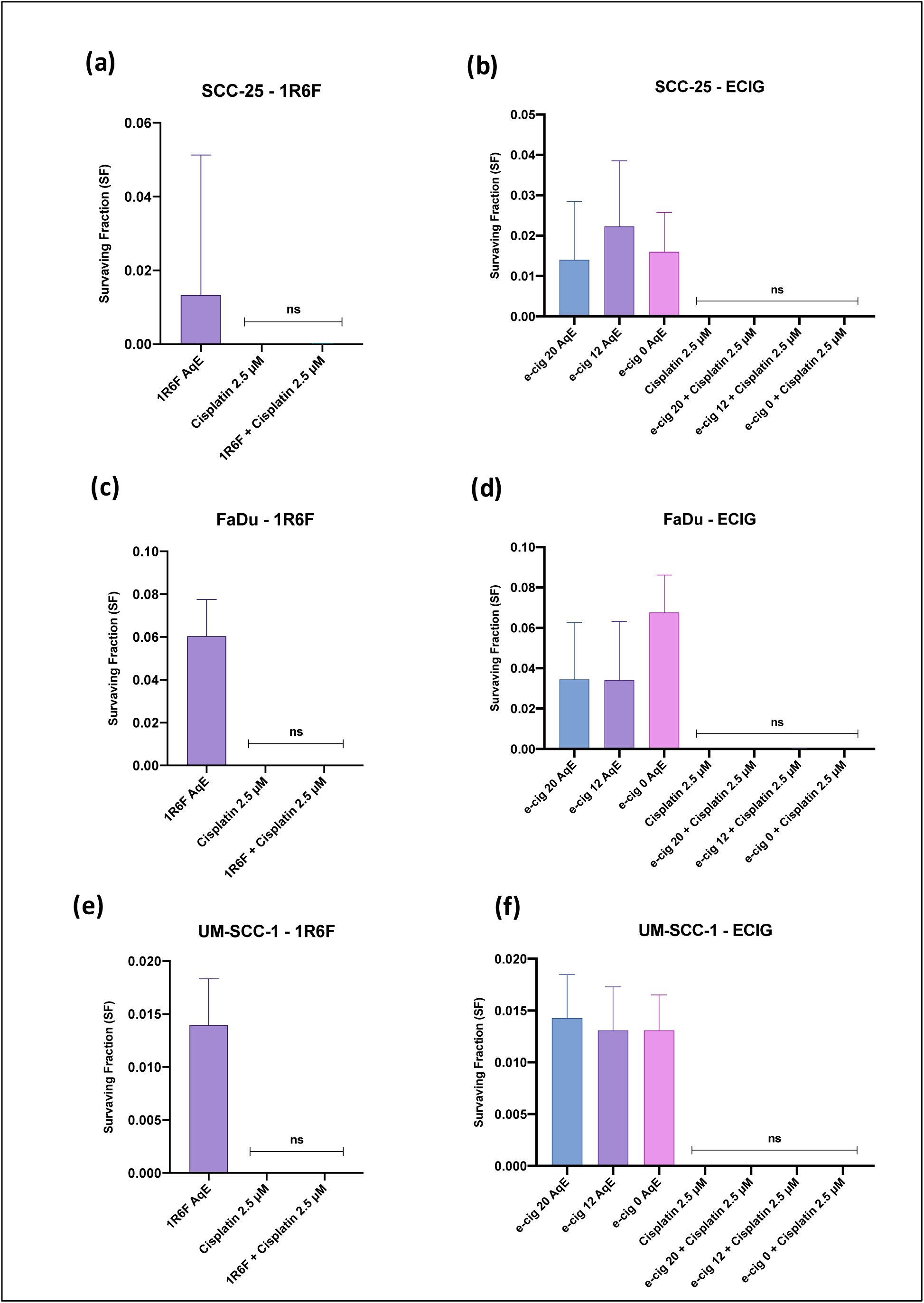
Clonogenic assay results for SCC-25, FaDu, and UM-SCC-1 cell lines. The survival fraction (SF) of each cell line was measured after treatment with 1R6F aqueous extract (AqE), cisplatin (2.5 µM), and combinations of e-cigarette extracts (20, 12, 0 AqE) with cisplatin. ns: not significant.

### mRNA expression of genes for the repair of cisplatin-induced DNA damage

We investigated whether cigarette and e-cigarette aerosol exposures affect the mRNA expression of XPA, MMS19, and ERCC1, which are critical genes for the repair of cisplatin-induced DNA damage. Compared to the original study, notable differences emerged in our results, particularly regarding the magnitude and direction of gene expression changes.

Across all cell lines (SCC-25, FaDu, UM-SCC-1), no significant differences were observed between treatments, indicating that XPA mRNA expression remained relatively stable irrespective of exposure condition. MMS19 mRNA expression was significantly downregulated only for SCC-25 following exposure to e-cig 12 nic compared to vehicle control (p= 0.047) and greater following exposure to e-cig 0 nic compared to control medium (p= 0.011) and vehicle control (p<0.0001). No significant differences in MMS19 mRNA expression were observed for FaDu and UM-SCC-1 cells. ERCC1 expression levels remained essentially unchanged in all cell lines and under all conditions, with only slight downregulation observed in SCC-25 for e-cig 0 compared to vehicle control alone (p= 0.039) and in FaDu for e-cig 20 compared to vehicle control alone (p= 0.012).

Our replication study agrees with Manyanga’s findings on the stability of XPA expression, but diverges significantly on the effects of e-cigarette aerosol on MMS19 and ERCC1. While Manyanga et al. (5) reported widespread and significant downregulation of MMS19 and ERCC1, our replication study found that these effects are sporadic and limited to specific conditions and cell lines, suggesting no nicotine dependent effect.

**Figure 7.**
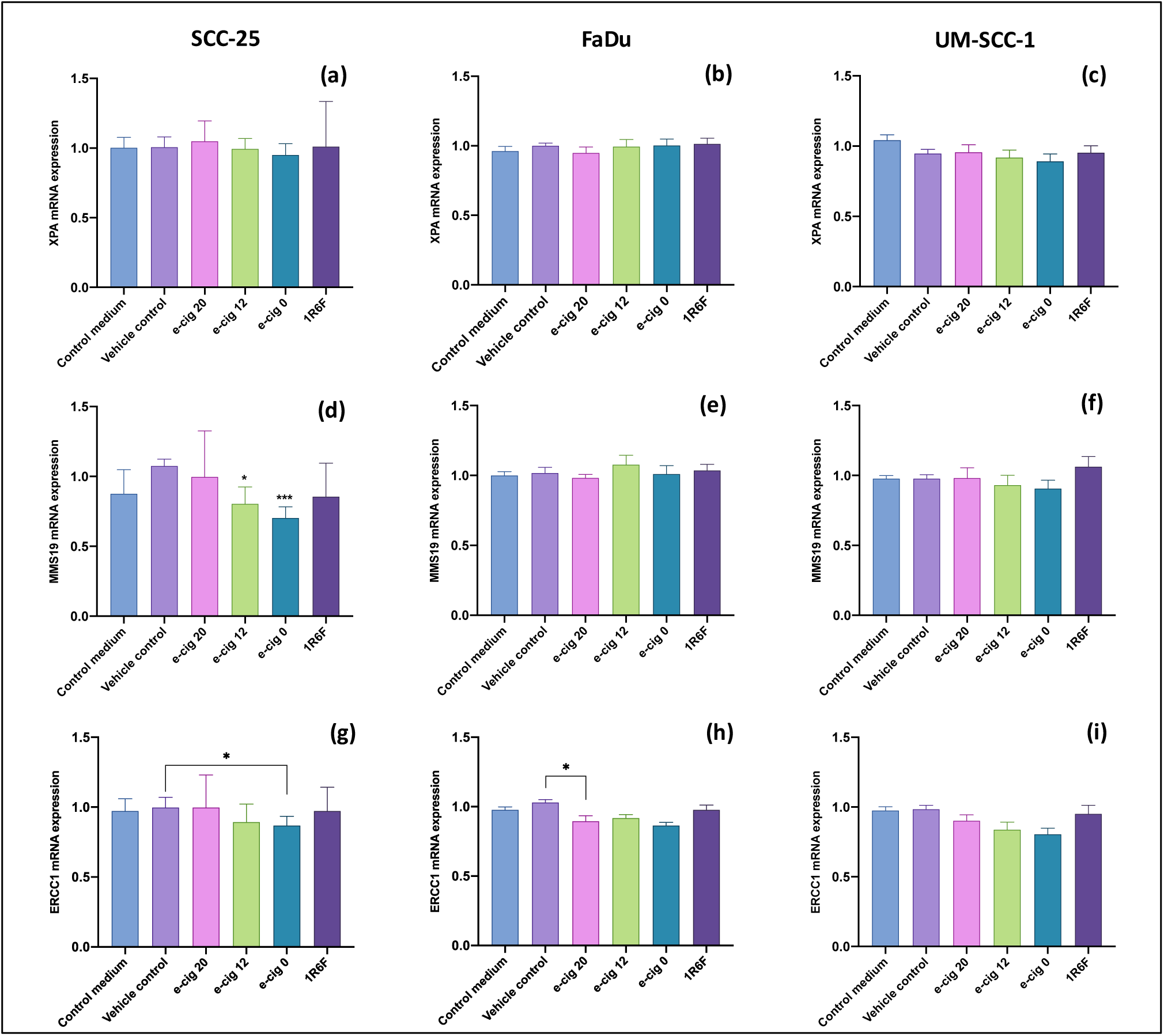
Expression of gene repair of cisplatin-induced DNA damage XPA (a, b, c), MMS19 (d, e f), and ERCC1 (g, h, i) in SCC-25, FaDu, and UM-SCC-1. Data in the panel (a), (d), and (g) were presented as median and CI 95% and were analyzed by using the Kruskal-Wallis test followed by Dunn’s test for multiple comparisons. Data in the panel (b), (c), (e), (f), (h), and (i) were presented as mean and SEM, and were analyzed by using ANOVA followed by Tukey’s test for multiple comparisons. *< 0.05; ***< 0.0001.

### Expression of drug influx and efflux transporter

#### mRNA expression results

We analyzed the mRNA expression of CTR1, ATP7A, ABCG2 (Fig. 8), ATP7B, ABCC2, ABCA1 and ABCC1 (Fig. 9) in SCC-25, FaDu and UM-SCC-1 cell lines following exposure to e-cigarette aerosol at various concentrations of nicotine (20, 12 and 0 mg/ml), 1R6F. The ATP7A gene expression assessment was performed by LAB-A and LAB-B in triplicate in two independent experimental sessions, except for the UM-SCC-1 with one experimental session by LAB-A and two experimental sessions by LAB-B. The gene expression assessment of CTR1, ABCG2, ATP7B, ABCC2, ABCA1, and ABCC1 was performed only by LAB-A in two independent experimental sessions for SCC-25 and FaDu, and one experimental session for UM-SCC-1. CTR1 expression levels remained stable for SCC-25 only, with no significant differences, but slight downregulation was observed for 1R6F compared to vehicle control in FaDu (p= 0.039) and for e-cig 20 nic compared to control medium in UM-SCC-1. Unlike Manyanga et al. (5) who reported significant upregulation of ATP7A in all cell lines analyzed, we observed no significant change in SCC-25, FaDu and UM-SCC-1. Evaluation of ABCG2 expression showed downregulation following exposure with e-cig 20 nic compared to controls in SCC-25 (p values < 0.05), e-cig 12 nic compared to control medium in FaDu (p= 0.021), and e-cig 0 nic compared to control medium in UM-SCC-1 (p= 0.01). This result contrasts with that reported by Manyanga et al. (5) who found significant upregulation of ABCG2 in all cell lines tested. ATP7B expression levels were found to be increased for e-cig 20 nic compared to controls only in SCC-25 (p values < 0.05) and decreased for e-cig 0 nic compared to control medium in UM-SCC-1. No significant changes were observed in FaDu. Unlike Manyanga’s results, the expression levels of ABCC2, ABCA1, and ABCC1 remained unchanged in all cell lines under all conditions.

**Figure 8.**
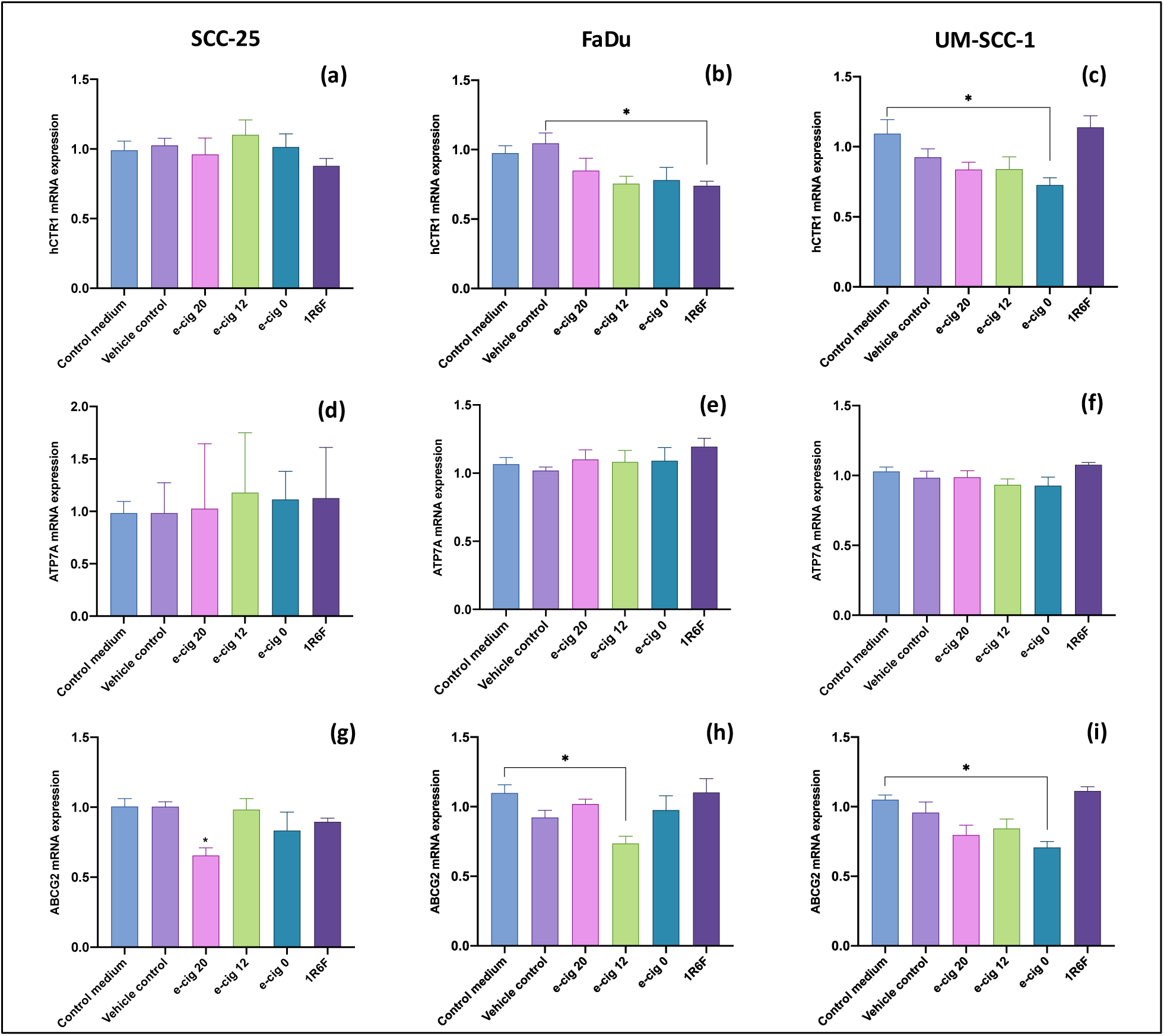
mRNA expression of drug influx and efflux transporter gene CTR1 (a, b, c), ATP7A (d, e f), and ABCG2 (g, h, i) in SCC-25, FaDu, and UM-SCC-1 cell lines. Data were presented as mean and SEM, and were analyzed using ANOVA followed by Tukey’s test for multiple comparisons. *< 0.05.

**Figure 9.**
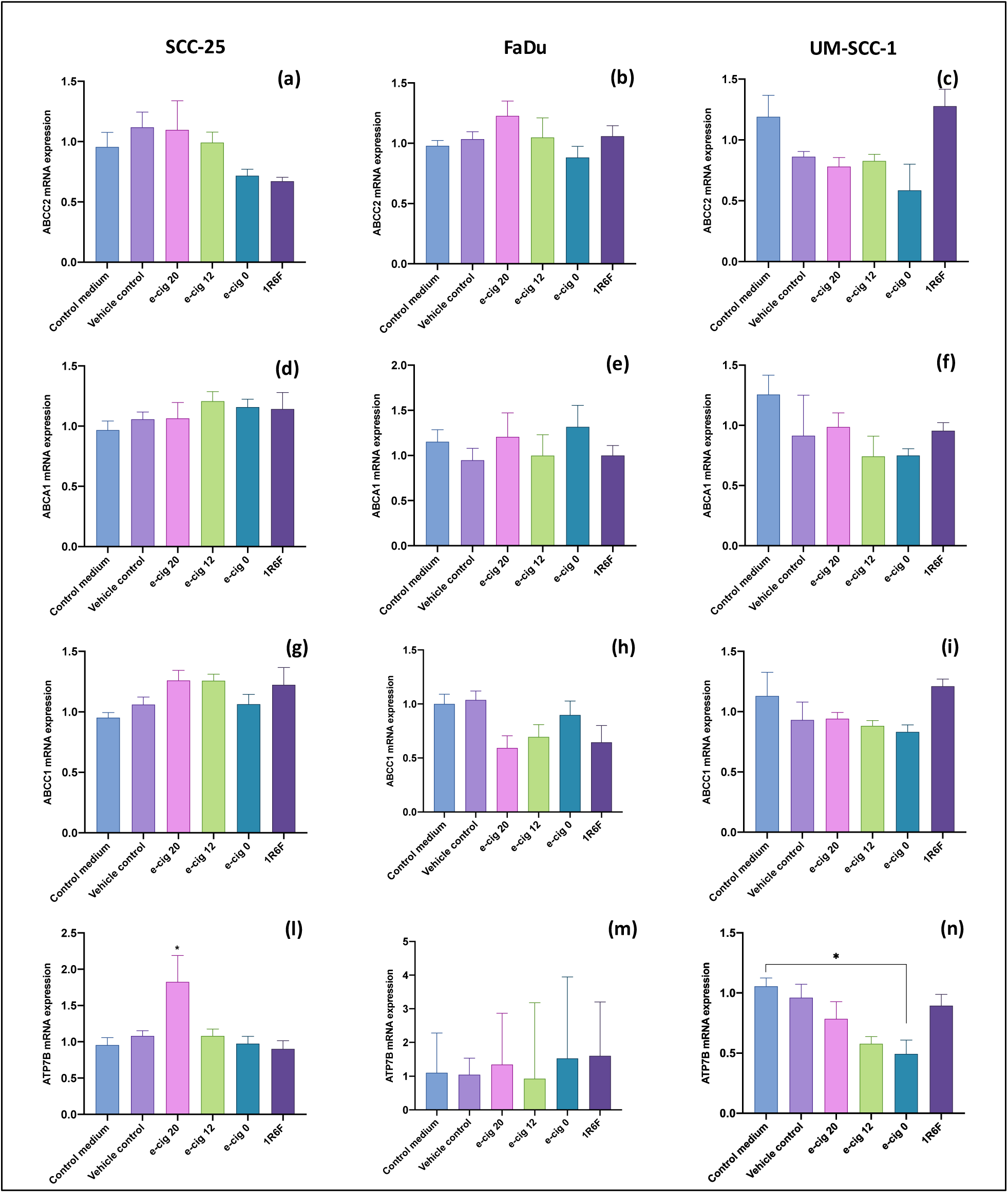
mRNA expression of drug influx and efflux transporter gene ABCC2 (a, b, c), ABCA1 (d, e f), ABCC1 (g, h, i), and ATP7B (l, m, n) in SCC-25, FaDu, and UM-SCC-1 cell lines. Data were presented as mean and SEM, and were analyzed using ANOVA followed by Tukey’s test for multiple comparisons. *< 0.05.

#### Western blot results

Western blot was performed by LAB-A, LAB-B, and LAB-D, but only LAB-A and LAB-D produced quantifiable results. As reported by Manyanga et al. (5), we investigated the protein expression of efflux (ATP7A and ABCG2) and influx (CTR1) transporter associated with cisplatin resistance in SCC-25, FaDu and UM-SCC-1 cell lines. Despite multiple attempts, none of the three proteins were detected in the pharyngeal cancer cell line (FaDu). Probably, these three proteins are poorly expressed in this cell line and their detection resulted to be difficult using the semi-dry method. For SCC-25 three independent experiments were performed by LAB-A and LAB-D. Whereas, two independent experiments for UM-SCC-1 were performed by LAB-A. All the unprocessed blot and the corresponding red ponceau are reported in figure S1, S2, and S3 of supplementary material. For the SCC-25 cells, LAB-A obtained protein expression results for ATP7A and CTR1 (Fig. S1); LAB-D obtained protein expression results for ATP7A, ABCG2 and CTR1 (Fig. S2). For UM-SCC-1 cells, LAB-A obtained protein expression results for ATP7A, ABCG2 and CTR1 (Fig. S3). For SCC-25, our results revealed no significant differences in protein expression among all experimental conditions. No significant alterations in ATP7A expression were observed between the control group and treatments with 1R6F, e-cigarette aerosol at 20, 12 or 0 mg/ml nicotine (Fig. 8a). ABCG2 expression levels were also unchanged in all treatment conditions (Fig. 8b). No significant changes in CTR1 protein expression were identified in any treatment condition (Fig. 8c). For UM-SCC-1, results showed selective changes in ATP7A protein expression (Fig. 8d). Notably, significant downregulation of ATP7A was observed in e-cigarette aerosol treatments compared with the control group. Specifically, e-cig 20 nic exposure led to moderate downregulation (p = 0.0207), while greater downregulation was observed with e-cig 12 nic (p = 0.0072) and e-cig 0 nic (p = 0.0032). ABCG2 expression levels were not significantly different in any of the conditions tested (Fig. 8e). CTR1 protein levels were also unchanged, with no significant differences between the control treatments (Fig. 8f).

Compared with Manyanga et al. (5), who reported consistent upregulation of ATP7A, downregulation of CTR1, and robust upregulation of ABCG2 in all cell lines, our study revealed distinct differences: ATP7A was significantly downregulated in UM-SCC-1 and unchanged in SCC-25, expression of CTR1 remained stable in both cell lines, and ABCG2 showed no significant changes in all conditions.

#### In cell western blot results

Protein expression levels of the influx transporter CTR1 and efflux transporters ATP7A and ABCG2 were also assessed by in-cell western blot analysis in SCC-25, FaDu, and UM-SCC-1 cell lines following exposure to e-cigarette aerosols (at 20, 12, and 0 mg/ml nicotine) and 1R6F conditions (Fig. 11). Evaluation by in cell western was performed only by LAB-C. The images of the plate used for quantification are included in the supplementary materials (Fig. S4-S12).

**Figure 10.**
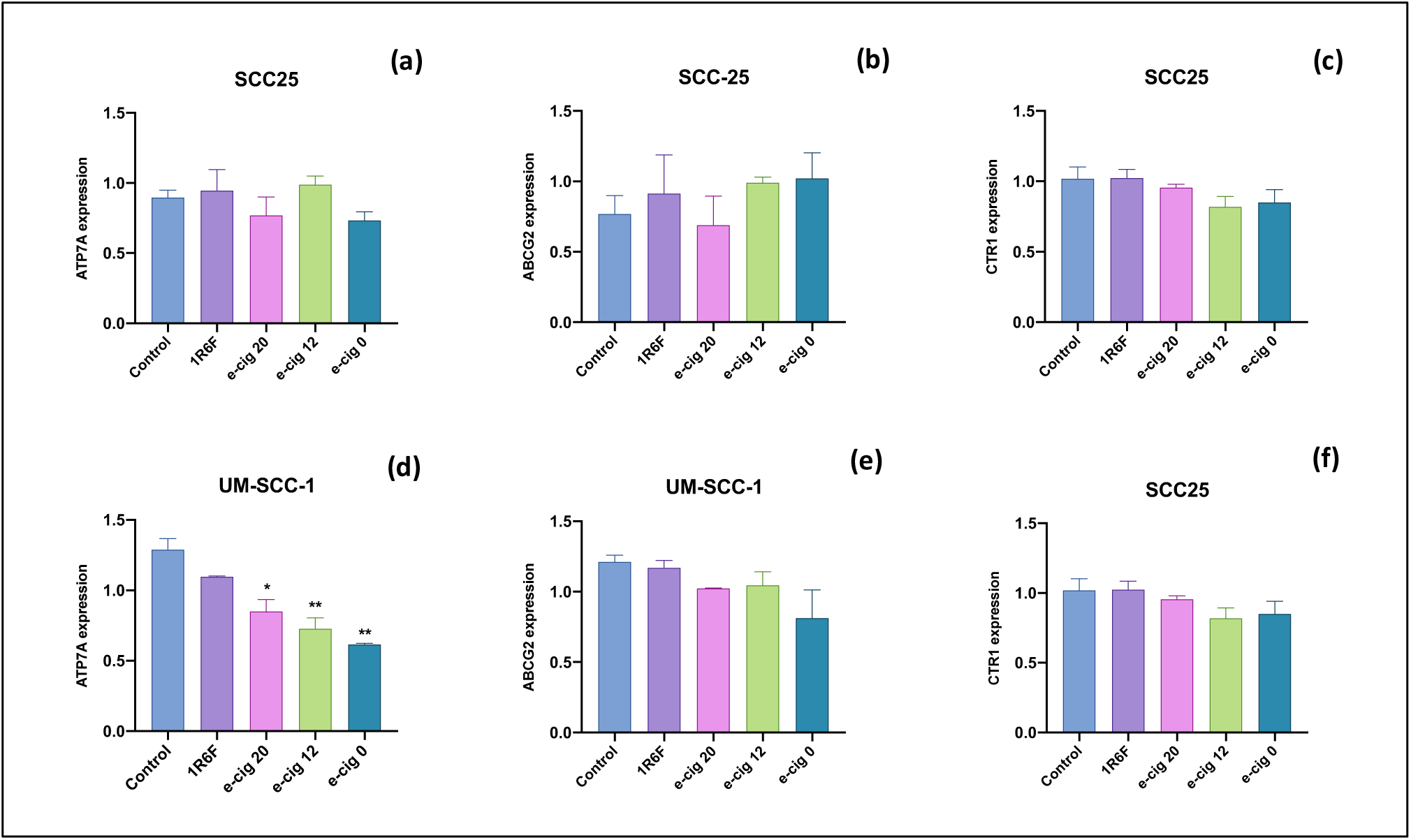
Protein expression of drug influx and efflux transporter in SCC-25 and UM-SCC-1 cells exposed to 1R6F and e-cigarette at different nicotine concentrations assessed by western blot. Data were presented as mean and SEM, and were analyzed used ANOVA followed by Tukey’s test for multiple comparisons. *< 0.05; **< 0.01.

**Figure 11.**
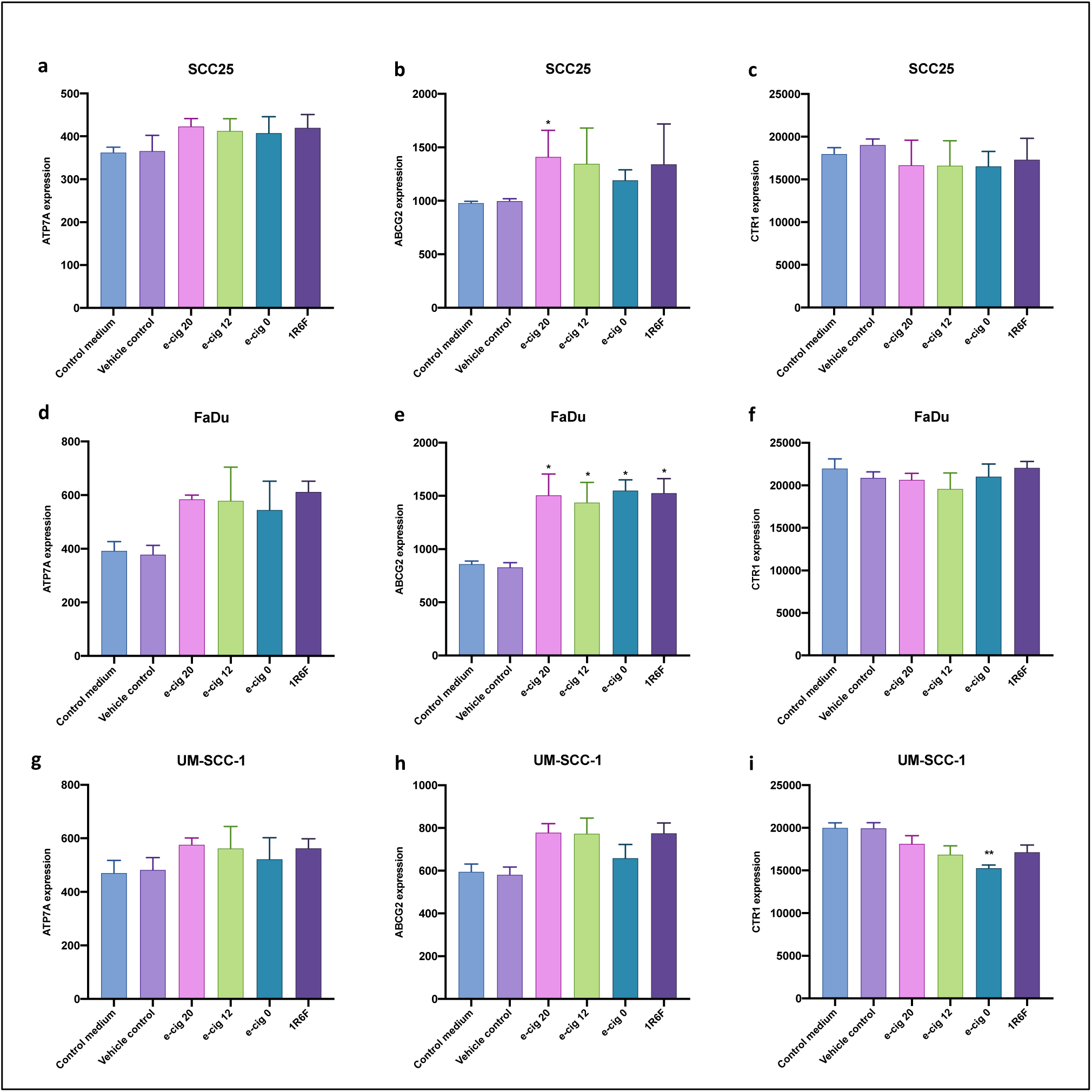
Protein expression of drug influx and efflux transporter in SCC-25 and UM-SCC-1 cells exposed to 1R6F and e-cigarette at different nicotine concentrations assessed by in-cell western blot. Data were presented as mean and SEM, and were analyzed using ANOVA followed by Tukey’s test for multiple comparisons. *< 0.05; **< 0.01.

ATP7A expression levels remained stable under all exposure conditions, with no significant differences between treatments in all three cell lines, indicating the stability of efflux transporter levels in this cell line. A significant increase in ABCG2 protein expression was observed in SCC-25s following e-cig 12 nic exposure compared with controls (p values < 0.05). In FaDu, ABCG2 expression was significantly increased in response to aerosols of 1R6F and e-cigarettes at all concentrations compared with controls (p values < 0.05). In addition, a significant increase was found following exposure to 1R6F (p < 0.05). In contrast, no significant changes in ABCG2 expression were observed for UM-SCC-1 under any treatment condition. No significant differences in CTR1 protein expression were observed in SCC-25 and FaDu under any of the experimental conditions. Expression levels remained constant in all treatments, including e-cigarette aerosols and 1R6F. In contrast, significant downregulation of CTR1 was noted in cells exposed to e-cigarette aerosol at 0 mg/ml nicotine compared with controls (p values < 0.01).

## Discussion

Our multicenter study aimed to replicate the study conducted by Manyanga et al. (5) on the effects of both cigarette smoke and e-cigarette aerosol on cisplatin resistance in head and neck cancer cell lines. Contrary to their observations, we found no significant modulation of cisplatin sensitivity in all three cell lines (SCC-25, FaDu, and UM-SCC-1) in response to exposure to e-cigarette aerosol and cigarette smoke (1R6F). The increase in colony formation by e-cigs and 1R6F observed in the original work was also not found in our results. However, we have partially replicated some results regarding the expression of genes and proteins associated with drug transporters and DNA repair pathways, but while Manyanga et al. (5) reported substantial alterations in gene and protein expression, our results indicate that these effects are sporadic and specific to the cell line and under only certain conditions tested.

Nicotine has been studied as a potential inducer of chemotherapy resistance, as it has been observed that smokers may show increased resistance to treatment in smokers (15). However, interactions between cancer therapies and smoking remain highly complex and not yet fully elucidated (16). Several papers have investigated how nicotine may influence cell survival and chemotherapy drug resistance through activation of specific signaling pathways and interaction with nicotinic receptors, emphasizing the importance of considering nicotine exposure in patients undergoing chemotherapy. Hsu et al. investigated the effect of nicotine on cell survival and cisplatin resistance in oral carcinoma by highlighting the role of α7-nicotinic acetylcholine receptors. The results showed that nicotine at concentrations ≥1 µM increases oral carcinoma cell survival, accelerates cell cycle progression and significantly reduces cisplatin-induced apoptosis (17). In another study, the regulation of cisplatin resistance in lung cancer cells by nicotine was explored. The results indicated that 1 µM nicotine increases cisplatin resistance through activation of specific signaling pathways (18). Other studies have also associated nicotine with increased chemotherapy resistance in several tumor types. In nasal carcinoma, it appears to reduce drug-induced apoptosis, impairing treatment efficacy (19). In lung carcinoma, it appears to modulate mitochondrial signaling and promote cell survival, as well as reduce Bcl-2 ubiquitination and degradation (20, 21). However, it is important to consider that most of these studies used high and unphysiologically relevant concentrations of nicotine for smokers and e-cigarette users. While concentrations above 1 µM are relevant for determining mechanistic effects *in vitro*, they do not necessarily reflect physiological exposures. Indeed, clinical studies reported nicotine plasma levels in smokers and vapers ranged from 0.025 to 0.44 µM (4 to 72 ng/ml) (22, 23). In their work, Manyanga et al. reported nicotine concentrations similar to that observed in the plasma of vapers (5.1 - 39 ng/ml / 0.031 µM - 0.241 µM) (5). Instead, our AqEs had nicotine concentrations between 0.44 and 0.63 µM (72 – 102.2 ng/ml). These concentrations are slightly higher than those in Manyanga et al.’s study, suggesting more intense or acute exposure to the compounds tested but still plausible for simulating normal to intense exposure scenarios. The main reason for the difference between the two studies lies in the standardization of AqE generation. Unlike Manyanga et al., in our study we used standardized smoking or vaping machines and regimens that provide greater control over experimental conditions. These differences in the exposure methods account for the different nicotine levels obtained between the two studies and underscore the importance of tightly controlled experimental conditions to ensure consistency and reproducibility of results. Beyond the effect of nicotine alone, it is important to note that cigarette smoke and e-cigarette aerosols are complex mixtures of different substances. Therefore, it is crucial to assess their health effects by considering the interactions among all aerosolized components, as Manyanga and colleagues have done, albeit with some limitations in the AqE generation that led to the substantial differences in our results.

In our study, the exposure to 1R6F or e-cigarette aerosol showed no significant impact on cisplatin-induced cytotoxicity in the three cell lines analyzed, as showed through MTS, NRU and Trypan Blue exclusion assays. Despite some observed variations, as in the case of Trypan Blue staining in UM-SCC-1 cells, the data were not statistically significant. This is in contrast to the results of Manyanga et al. who showed an increase in cisplatin resistance in the presence of e-cigarette aerosol. Like Manyanga’s work, Simon and colleagues observed that cigarette smoke condensate reduced the efficacy of cisplatin in head and neck cancer cells (PiCa and FaDu), showing increased cell viability and proliferation. However, this increase was not very pronounced and, as in our work, they did not observe an increase in cell viability for FaDu cells (24). Moreover, the evaluation of the IC50 values of cisplatin obtained in combination with 1R6F and e-cigarette aerosol was not replicated in our work. Overall, we did not observe a reduction in cisplatin sensitivity in the three head and neck cancer cell lines. For the FaDu line, exposure to 1R6F resulted in increased sensitivity to cisplatin, with significantly lower IC50 values, suggesting a more sensitive response to the combination. However, this could be due to the cytotoxic effect of the combination of cisplatin and combustion byproducts. Also, exposure to e-cigarette aerosol did not produce significant changes in IC50 value in all cell lines except for the MTS evaluation in UM-SCC-1, where a significant decrease in IC50 values was observed compared to cisplatin alone. As observed in Manyanga studies (5, 25), we confirmed that neither e-cigarette extracts alone, nor corresponding nicotine concentrations increased cell proliferation. In fact, both nicotine-containing and nicotine-free e-cigarette extracts produced comparable effects on cisplatin-induced cell death and IC50 values. These results contrast with previous studies that reported increased tumor cell proliferation in response to nicotine exposure (17, 26). The use of the NRU test in our work deserves special recognition, as it increases the consistency of results in our multicenter study. This test is often used in the evaluation of tobacco products because of its greater sensitivity in detecting cell viability than other commonly used viability tests (27, 28). Moreover, our results showed high reproducibility in cytotoxicity and chemosensitivity assays, with better performance of NRU than MTS. This high reproducibility is particularly important to emphasize the reliability of these results.

The clonogenic survival assay is a widely used method to assess the ability of cells to proliferate and form colonies after exposure to harmful treatments, such as chemotherapeutic agents or exposure to toxic substances. This technique provides information on the reproductive capacity of cells and is a key indicator of resistance or sensitivity to anticancer treatments (14). In our study, the clonogenic assay was used to analyze the effect of cisplatin in combination with traditional cigarette (1R6F) and e-cigarette extracts on three HNSCC cell lines. Consistent with the results obtained from cytotoxicity and chemosensitivity assays, the clonogenic results indicate that, in all three cell lines, there was no significant interaction between cisplatin alone and cisplatin combined with 1R6F extracts and e-cigarette aerosol. Furthermore, exposure to 1R6F and e-cigarette aerosol extracts, in the absence of cisplatin, did not significantly alter the clonogenic capacity of the cells. These results differ from those reported by Manyanga et al. (5), who observed that nicotine in e-cigarette aerosols can increase clonogenic survival after cisplatin treatment in HNSCC cell lines. In our study, no such effect was found, even for the UM-SCC-1 line derived from a smoking patient. Unfortunately, there are no other data in the literature investigating the effects of e-cigarette aerosols on colony-forming ability in combination with cisplatin. Our results suggest that the use of e-cigarettes during cisplatin chemotherapy may not increase the risk of clonogenic survival of HNSCC tumor cells. The clonogenic assay reproducibility results indicated that our data are reasonably consistent among the cell lines, particularly for FaDu and UM-SCC-1, where moderate to high reproducibility was observed. While the reproducibility for SCC-25 was more limited, the statistical significance of the ICC values supports the reliability of our results. This reinforces the validity of our conclusions and underscores scores the robustness of our experimental approach. However, discrepancies with Manyanga’s results underscore the need for further studies to clarify the effect of nicotine and other components of e-cigarette aerosols on different tumor lines and under varying experimental conditions.

Cisplatin is an intercalating agent known for its ability to bind to purine bases in DNA, generating damage that interferes with repair mechanisms and inducing cell cycle arrest and apoptosis (29). However, cisplatin resistance is a complex phenomenon, mediated by genetic and epigenetic modifications involving several pathways, including those for DNA repair (30). Replicating Manyanga’s study (5), we evaluated whether exposure to aerosols from cigarettes and e-cigarettes affects the expression of three genes critical for cisplatin-induced DNA damage repair: XPA, MMS19 and ERCC1. Again, our results showed significant differences from Manyanga et al.’s work (5), especially regarding the magnitude and direction of changes in gene expression, particularly for MMS19 and ERCC1. Our results showed sporadic downregulation of MMS19, limited to the SCC-25 cell line and independent of nicotine concentration, and slight downregulation of ERCC1 under some conditions (SCC-25 exposed to e-cig 0 nic and in FaDu with e-cig 20 nic), but globally stable. Consistent with the results of Manyanga et al (5), XPA expression was not significantly altered under any of the conditions tested. This finding confirms that the expression of this gene remains stable, even with exposure to cigarette aerosols and e-cigarettes, suggesting that the role of XPA in cisplatin-induced DNA repair may not be directly modulated by these treatments. Unlike Manyanga et al. who reported more pronounced and widespread effects, our results suggest a less uniform influence of aerosols on DNA repair, with variations dependent on cell line and type of exposure. Variability remains a critical factor in reproducibility data, especially for cell lines such as FaDu and UM-SCC-1. However, although reproducibility is not ideal, results showing good to moderate concordance for MMS19 and ERCC1 gene expression are indicative of higher reliability. Moreover, given our rigorous and standardized approach to reduce systematic experimental errors, the variability observed in the FaDu and UM-SCC-1 cell lines could be due to their intrinsic biological response to aerosols, rather than methodological flaws (31).

Membrane transporters play a crucial role in determining the efficacy of cisplatin therapy, influencing both the entry and efflux of the drug from tumor cells. In this study, as previously done by Manyanga and colleagues (5), we analyzed the expression of several key transporters involved in cisplatin resistance: CTR1 represents the main active transport system for cisplatin entrance into cells, and its expression is directly correlated with intracellular accumulation of the drug and, consequently, with the efficacy of therapy (32, 33); the efflux transporters ATP7A and ATP7B are P-type ATPases involved in the transport of copper and platinum. In particular, ATP7A plays a key role in the expulsion of cisplatin from cells, and its increased expression has been correlated with reduced response to therapy (34); the ABC (ATP-Binding Cassette) family of transporters includes ABCG2, ABCC1, ABCC2, and ABCA1, which contribute to multidrug resistance through ATP-dependent transport of various chemical compounds (35). Among them, ABCG2 seems to be involved in cisplatin resistance associated with tobacco smoke exposure (24). Regarding mRNA expression of membrane transporters, our study revealed substantially different results from Manyanga et al. While they observed significant upregulation of ATP7A in all cell lines, we found no significant changes in the expression of this gene in either SCC-25, FaDu or UM-SCC-1 cells. For ABCG2, our results show downregulation in all cell lines, in contrast to the significant upregulation reported by Manyanga(5). Furthermore, unlike Manyanga et al., which observed significant increases in ABCC2, ABCA1 and ABCC1 in at least one cell line, our results showed no alterations in the expression of these transporters under any of the conditions tested. Western blot results showed fundamental differences from Manyanga results. ATP7A was downregulated in UM-SCC-1 with e-cigarette treatment and unchanged in SCC-25. CTR1 remained stable in both cell lines as well as ABCG2 expression showed no change, in contrast to the upregulation reported by Manyanga for all the three transporters. Also, in-cell Western blot results performed by LAB-C showed variability among cell lines. Only ABCG2 was upregulated in SCC-25 for e-cig 20 nic and FaDu for all e-cigs but not in UM-SCC-1. Meanwhile, CTR1 was downregulated only for e-cig 0 nic in UM-SCC-1. Our results highlight the variability in transporter regulation among cell lines and suggest that cisplatin resistance is not entirely determined by environmental factors, such as exposure to e-cigarette components. This indicates the need to consider intrinsic cellular factors, such as genetic or metabolic differences, to understand and overcome cisplatin resistance.

The strengths of this study lie in its rigorous multicenter design and harmonization of experimental protocols among different international laboratories, which increases the consistency of our results. The use of technical triplicates and multiple experimental sessions contribute to the robustness of the data. In addition, by using standardized operating procedures (SOPs), our research ensured consistency in the experimental process. However, some limitations should be considered. The late acquisition of the UM-SCC-1 cell line by LAB-A resulted in experiments conducted in a single session, reducing the data sample analyzed. The limited application of the in-cell western assay by only one laboratory also reduces the consistency of these results. The replacement of WSU-HN6 and WSU-HN30 cell lines with other alternatives may limit direct comparison with Manyanga’s study. Finally, acute *in vitro* exposure may not be representative of the more complex long-term exposure in humans.

In conclusion, several of the findings reported by Manyanga et al (5) regarding the ability of e-cigarette aerosol to promote cisplatin resistance in head and neck cancer cell lines, were not confirmed in our multicenter study. Although we observed some effects on gene expression, these were sporadic and observed on specific cell lines under certain exposure conditions. Therefore, our results suggest that nicotine and e-cigarette aerosol components do not consistently influence cisplatin resistance, and the variability observed in gene expression underscores the importance of considering cell-specific responses when assessing the impact of these substances on cancer treatment outcomes. In addition, our study highlights the need for standardized protocols in aerosol exposure research to ensure reproducibility and reliability of results. Although our study did not replicate the increase in cisplatin resistance observed in previous studies, it contributes valuable data to the ongoing debate about the nicotine use and cancer therapy, suggesting that more comprehensive investigations are needed to better understand these effects and their clinical implications.

## Statements and Declarations

### Data availability

The datasets used and/or analyzed during the current study are available from the corresponding author on reasonable request.

