## Supplementary Materials for "Multi-Center “Replica Study” Challenges the Impact of Electronic Cigarette Aerosols on Cisplatin Resistance in Head and Neck Cancer Cells"

### Nicotine quantification

Nicotine in AqE was analyzed by reversed-phase HPLC. The system included a SpectraSystem P4000 pump coupled to a UV6000LP diode array detector (ThermoQuest Italy), with a 5-cm flow cell operating at 190-300 nm. The different AqE samples were analyzed on a Hypersil Gold RP C18 column (150 × 4.6 mm, 3 µm) with a guard column (Thermo Fisher Scientific). Separation used a binary gradient of buffer A (10% methanol, 90% 10mM $KH_2PO_4$, pH 7.4) and buffer B (50% methanol, 50% 10mM $KH_2PO_4$, pH 7.4) programmed as: 0-5 min 100% A, 5-30 min 100% B. The column was maintained at 30°C with a flow rate of 1 ml/min. Nicotine eluted at k' = 13 (k' = V - V0/V0, where V = elution volume of the compound, V0 = void volume of the system) and was quantified at its maximum absorbance (λ= 268 nm).

**Table S1. Primer sequences used for RT-PCR.**

| Product | Nicotine in the product | Nicotine in the 0.2 puffs/ml AqE stock (µM) | Nicotine in the 0.002 puffs/ml work solution (µM) |
|---|---|---|---|
| **1R6F** | 1.896 mg/cigarette* | 44.15 | 0.44 |
| **e-cig 20** | 20 mg/ml | 63.22 | 0.63 |
| **e-cig 12** | 12 mg/ml | 46.40 | 0.46 |
| **e-cig 0** | 0 mg/ml | 0.00 | 0.00 |
| *following the HCI regime | | | |

### Gene expression by Real-Time PCR

**Table S2. Primer sequences used for RT-PCR.**

| ***Primer** | **Forward** | **Reverse** |
|---|---|---|
| β-actin | 5'-GTCATCACCATTGGCAATGAG-3' | 5'-ATGTCCACGTCACACTTCATG-3' |
| ATP7A | 5'-GCAGAGCCTCTATAAACTCAC-3’ | 5’-GTGTCATCATCTTCCCTGAAG-3' |
| ATP7B | 5'-GACCAGGTCAGCTATGTCAG-3’ | 5’-CATCAGATGTACTGCTCCTCA-3' |
| hCTR1 | 5'-CTCATCTTCATGACCTACAACG-3’ | 5’-GATGTCAATGGCAATGCTCTG-3' |
| ABCG2 | 5'-TGTGGCATTAAACAGAGAAGAAGAC-3' | 5'-TCACCCCCGGAAAGTTGATG-3' |
| ABCA1 | 5'-CTCAGACAACACTTGACCAAG-3’ | 5’-AGATGTGAGAACTGCAACGTC-3' |
| ABCC1 | 5'-ACACAGTTCGAGGACTGCAC-3’ | 5’-GAAAAGACCTCTCTGCTGCAG-3' |
| ABCC2 | 5'-AGGTAATGGTCCTAGACAACG-3’ | 5’-GTGCTGTTCACATTCTCAATGC-3' |
| ERCC1 | 5’ -GGCGACGTAATTCCCGACT-3’ | 5’ -TAGCGGAGGCTGAGGAACA-3’ |
| XPA | 5'-GCAGCCCCAAAGATAATTGA-3' | 5'-TGGCAAATCAAAGTGGTTCA-3' |
| MMS19 | 5'-GTCAGCAGGACCCTGAGAGTTC-3' | 5'-CTCAGAACTGAGGGCTCCTTC-3' |
| **Primer details: HPSF - salt free - 0.01** | | |

### Reproducibility across all laboratories

**Table S3.** Cell viability results of ICC Calculation using absolute-agreement, two-way mixed-effects model.

| | Raters (LAB) | ICC | 95% Confidence interval | | F-Test with true value 0 | | | |
|---|---|---|---|---|---|---|---|---|
| | | | *Lower bound* | *Upper bound* | *Value* | *Df1* | *Df2* | *p value* |
| **MTS SCC-25 1R6F** | 6 (A;B;C;D) | 0.884 | 0.592 | 0.99 | 63.6 | 2 | 13.3 | <0,0001 |
| **MTS SCC-25 ECIG** | 8 (A;B;C;D) | 0.556 | 0.257 | 0.871 | 25.7 | 6 | 16.8 | <0,0001 |
| **MTS FaDu 1R6F** | 6 (A;B;C) | 0.609 | 0.184 | 0.985 | 19.6 | 2 | 10.7 | 0.0003 |
| **MTS FaDu ECIG** | 5 (A;B;C) | 0.506 | 0.144 | 0.858 | 18.9 | 6 | 7.63 | 0.0003 |
| **MTS UM-SCC-1 1R6F** | 5 (A;B;C) | 0.598 | 0.147 | 0.984 | 13.4 | 2 | 9.81 | 0.0016 |
| **MTS UM-SCC-1 ECIG** | 5 (A;B;C) | 0.574 | 0.253 | 0.883 | 10.3 | 6 | 21 | <0,0001 |
| **NRU SCC-25 1R6F** | 8 (A;B;C;D) | 0.665 | 0.261 | 0.988 | 46.4 | 2 | 11.8 | <0,0001 |
| **NRU SCC-25 ECIG** | 8 (A;B;C;D) | 0.646 | 0.373 | 0.905 | 20.9 | 6 | 36 | <0,0001 |
| **NRU FaDu 1R6F** | 6 (A;B;C) | 0.932 | 0.729 | 0.998 | 109 | 2 | 13.5 | <0,0001 |
| **NRU FaDu ECIG** | 6 (A;B;C) | 0.906 | 0.725 | 0.981 | 122 | 6 | 13.4 | <0,0001 |
| **NRU UM-SCC-1 1R6F** | 3 (A;B;C) | 0.616 | 0.026 | 0.985 | 14.2 | 2 | 3.72 | <0,0001 |
| **NRU UM-SCC-1 ECIG** | 4 (A;B;C) | 0.5 | 0.141 | 0.857 | 7.94 | 6 | 12.1 | <0,0001 |

The table includes the ICC values, 95% confidence intervals, F-test values, degrees of freedom (Df1 and Df2), and p-values for each group of raters.

**Table S4.** Trypan-blue results of ICC Calculation using absolute-agreement, two-way mixed-effects model.

| | Raters (LAB) | ICC | 95% Confidence interval | | F-Test with true value 0 | | | |
|---|---|---|---|---|---|---|---|---|
| | | | *Lower bound* | *Upper bound* | *Value* | *Df1* | *Df2* | *p value* |
| **SCC-25 1R6F** | 8 (A;B;C;D) | 0.154 | 0.031 | 0.467 | 4.07 | 8 | 30.3 | 0.0022 |
| **SCC-25 ECIG** | 8 (A;B;C;D) | 0.119 | 0.034 | 0.277 | 3.65 | 20 | 33.1 | 0.0005 |
| **FaDu 1R6F** | 6 (A;B;C) | 0.534 | 0.268 | 0.827 | 9.5 | 8 | 37.4 | <0,0001 |
| **FaDu ECIG** | 6 (A;B;C) | 0.648 | 0.468 | 0.81 | 15.2 | 20 | 57.8 | <0,0001 |
| **UM-SCC-1 1R6F** | 3 (A;B;C) | 0.634 | 0.257 | 0.888 | 7.9 | 8 | 13.6 | 0.0005 |
| **UM-SCC-1 ECIG** | 3 (A;B;C) | 0.826 | 0.625 | 0.925 | 21.1 | 20 | 16.8 | <0,0001 |

The table includes the ICC values, 95% confidence intervals, F-test values, degrees of freedom (Df1 and Df2), and p-values for each group of raters.

**Table S5.** Chemosensitivity results of ICC Calculation using absolute-agreement, two-way mixed-effects model.

| | | | **95% Confidence interval** | | **F-Test with true value 0** | | | |
|---|---|---|---|---|---|---|---|---|
| | **Raters (LAB)** | **ICC** | ***Lower bound*** | ***Upper bound*** | ***Value*** | ***Df1*** | ***Df2*** | ***p value*** |
| **MTS SCC-25 1R6F** | 6 (A;B;C;D) | 0.943 | 0.868 | 0.983 | 127 | 9 | 36 | <0.0001 |
| **MTS SCC-25 ECIG** | 8 (A;B;C;D) | 0.895 | 0.81 | 0.951 | 94.4 | 19 | 59 | <0.0001 |
| **MTS FaDu 1R6F** | 6 (A;B;C) | 0.899 | 0.778 | 0.969 | 68.7 | 9 | 36.3 | <0.0001 |
| **MTS FaDu ECIG** | 5 (A;B;C) | 0.882 | 0.781 | 0.946 | 48.2 | 19 | 44.6 | <0.0001 |
| **MTS UM-SCC-1 1R6F** | 5 (A;B;C) | 0.867 | 0.714 | 0.959 | 38.5 | 9 | 35 | <0.0001 |
| **MTS UM-SCC-1 ECIG** | 5 (A;B;C) | 0.796 | 0.655 | 0.901 | 23.9 | 19 | 58.7 | <0.0001 |
| **NRU SCC-25 1R6F** | 8 (A;B;C;D) | 0.896 | 0.785 | 0.968 | 75.1 | 9 | 66.9 | <0.0001 |
| **NRU SCC-25 ECIG** | 8 (A;B;C;D) | 0.867 | 0.773 | 0.936 | 64.7 | 19 | 87.7 | <0.0001 |
| **NRU FaDu 1R6F** | 6 (A;B;C) | 0.942 | 0.869 | 0.983 | 113 | 9 | 43.9 | <0.0001 |
| **NRU FaDu ECIG** | 6 (A;B;C) | 0.949 | 0.905 | 0.976 | 129 | 19 | 73.6 | <0.0001 |
| **NRU UM-SCC-1 1R6F** | 3 (A;B;C) | 0.885 | 0.685 | 0.968 | 32 | 9 | 13.2 | <0.0001 |
| **NRU UM-SCC-1 ECIG** | 4 (A;B;C) | 0.894 | 0.804 | 0.951 | 37.9 | 19 | 52.9 | <0.0001 |

The table includes the ICC values, 95% confidence intervals, F-test values, degrees of freedom (Df1 and Df2), and p-values for each group of raters.

**Table S6.** Clonogenic results of ICC Calculation using absolute-agreement, two-way mixed-effects model.

| | | | **95% Confidence interval** | | **F-Test with true value 0** | | | |
|---|---|---|---|---|---|---|---|---|
| | **Raters (LAB)** | **ICC** | ***Lower bound*** | ***Upper bound*** | ***Value*** | ***Df1*** | ***Df2*** | ***p value*** |
| **SCC-25 1R6F** | 8 (A;B;C;D) | 0.154 | 0.031 | 0.467 | 4.07 | 8 | 30.3 | 0.0022 |
| **SCC-25 ECIG** | 8 (A;B;C;D) | 0.119 | 0.034 | 0.277 | 3.65 | 20 | 33.1 | 0.0005 |
| **FaDu 1R6F** | 6 (A;B;C) | 0.534 | 0.268 | 0.827 | 9.5 | 8 | 37.4 | <0,0001 |
| **FaDu ECIG** | 6 (A;B;C) | 0.648 | 0.468 | 0.81 | 15.2 | 20 | 57.8 | <0,0001 |
| **UM-SCC-1 1R6F** | 3 (A;B;C) | 0.634 | 0.257 | 0.888 | 7.9 | 8 | 13.6 | 0.0005 |
| **UM-SCC-1 ECIG** | 3 (A;B;C) | 0.826 | 0.625 | 0.925 | 21.1 | 20 | 16.8 | <0,0001 |

The table includes the ICC values, 95% confidence intervals, F-test values, degrees of freedom (Df1 and Df2), and p-values for each group of raters.

**Table S7.** Gene expression results of ICC Calculation using absolute-agreement, two-way mixed-effects model.

| | Raters (LAB) | ICC | 95% Confidence interval | | F-Test with true value 0 | | | |
|---|---|---|---|---|---|---|---|---|
| | | | *Lower bound* | *Upper bound* | *Value* | *Df1* | *Df2* | *p value* |
| **SCC-25 XPA** | 8 (A;B;C;D) | 0.0163 | -0.078 | 0.418 | 1.14 | 5 | 36.7 | 0.357 |
| **SCC-25 MMS19** | 8 (A;B;C;D) | 0.36 | 0.115 | 0.798 | 7.32 | 5 | 33.8 | <0.0001 |
| **SCC-25 ERCC1** | 8 (A;B;C;D) | 0.171 | 0.006 | 0.64 | 3.09 | 5 | 40.8 | 0.0187 |
| **SCC-25 ATP7A** | 4 (A;B) | 0.0466 | -0.103 | 0.531 | 1.32 | 5 | 17.7 | 0.301 |
| **FaDu XPA** | 6 (A;B;C) | -0.0775 | -0.132 | 0.201 | 0.438 | 5 | 7.79 | 0.811 |
| **FaDu MMS19** | 6 (A;B;C) | -0.0799 | -0.131 | 0.187 | 0.41 | 5 | 6.58 | 0.828 |
| **FaDu ERCC1** | 6 (A;B;C) | 0.283 | 0.017 | 0.77 | 3.35 | 5 | 29.4 | 0.0162 |
| **FaDu ATP7A** | 4 (A;B) | -0.0205 | -0.075 | 0.255 | 0.759 | 5 | 8.46 | 0.602 |
| **UM-SCC-1 XPA** | 5 (A;B;C) | 0.0494 | -0.058 | 0.467 | 1.5 | 5 | 23.9 | 0.228 |
| **UM-SCC-1 MMS19** | 5 (A;B;C) | -0.0105 | -0.071 | 0.293 | 0.884 | 5 | 16.6 | 0.514 |
| **UM-SCC-1 ERCC1** | 5 (A;B;C) | 0.151 | -0.019 | 0.627 | 2.73 | 5 | 18.9 | 0.0511 |
| **UM-SCC-1 ATP7A** | 3 (A;B) | 0.0286 | -0.194 | 0.592 | 1.13 | 5 | 10.9 | 0.4 |

The table includes the ICC values, 95% confidence intervals, F-test values, degrees of freedom (Df1 and Df2), and p-values for each group of raters.

**Table S8.** Western Blot results of ICC Calculation using absolute-agreement, two-way mixed-effects model.

| | Raters (LAB) | ICC | 95% Confidence interval | | F-Test with true value 0 | | | |
|---|---|---|---|---|---|---|---|---|
| | | | *Lower bound* | *Upper bound* | *Value* | *Df1* | *Df2* | *p value* |
| **SCC-25 ATP7A** | 8 (A;B;C;D) | 0.132 | -0.03 | 0.672 | 2.58 | 4 | 16.7 | 0.0751 |
| **SCC-25 ABCG2** | 8 (A;B;C;D) | -0.173 | -0.465 | 0.603 | 0.58 | 4 | 6.19 | 0.689 |
| **SCC-25 CTR1** | 6 (A;B;C) | 0.12 | -0.083 | 0.706 | 1.82 | 4 | 23.3 | 0.159 |

The table includes the ICC values, 95% confidence intervals, F-test values, degrees of freedom (Df1 and Df2), and p-values for each group of raters.

**Western blot: unprocessed blot and the corresponding red ponceau**

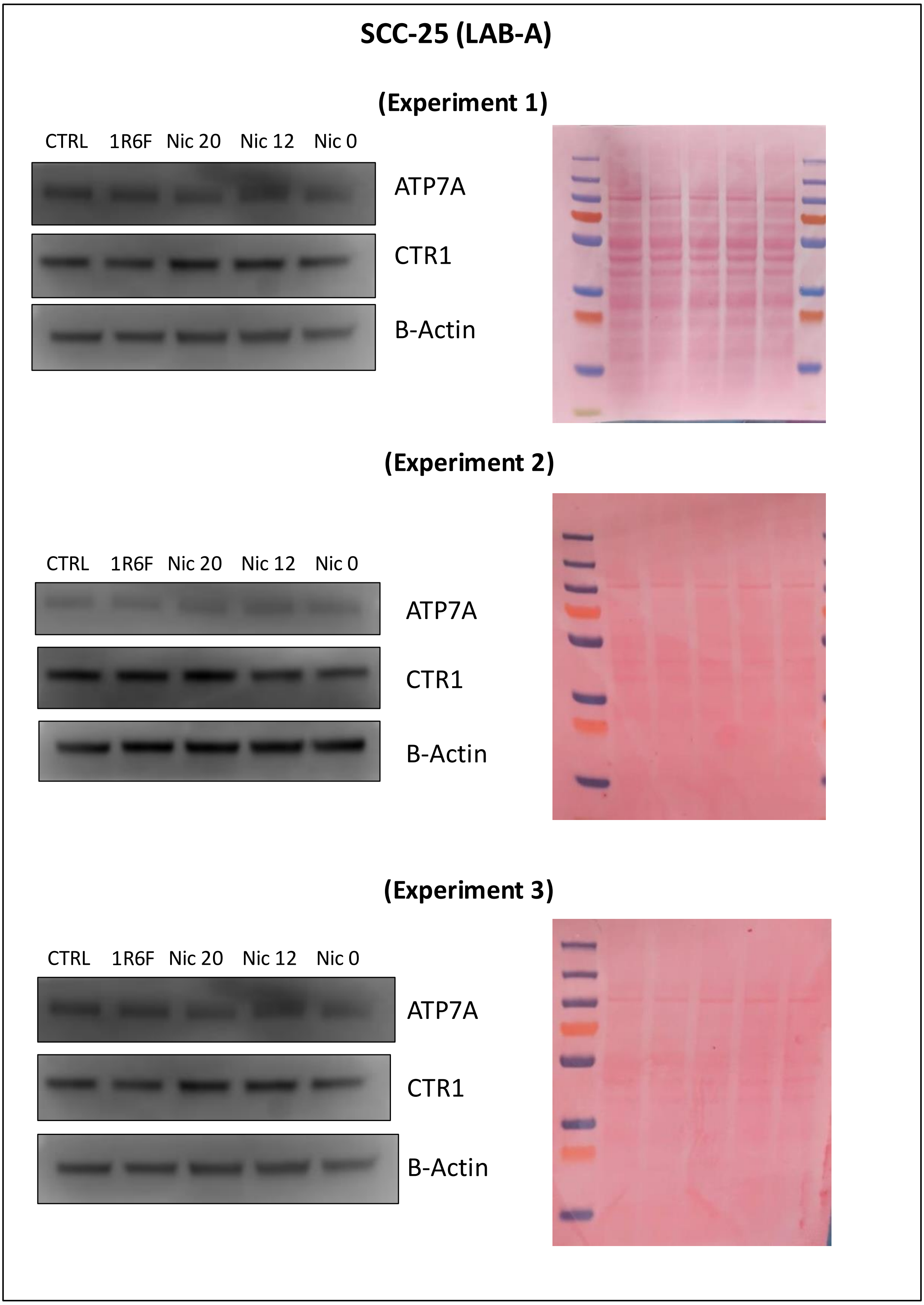


**Figure S1.** Unprocessed blot and the corresponding red ponceau of western blot in SCC-25 cell line executed by LAB-A.

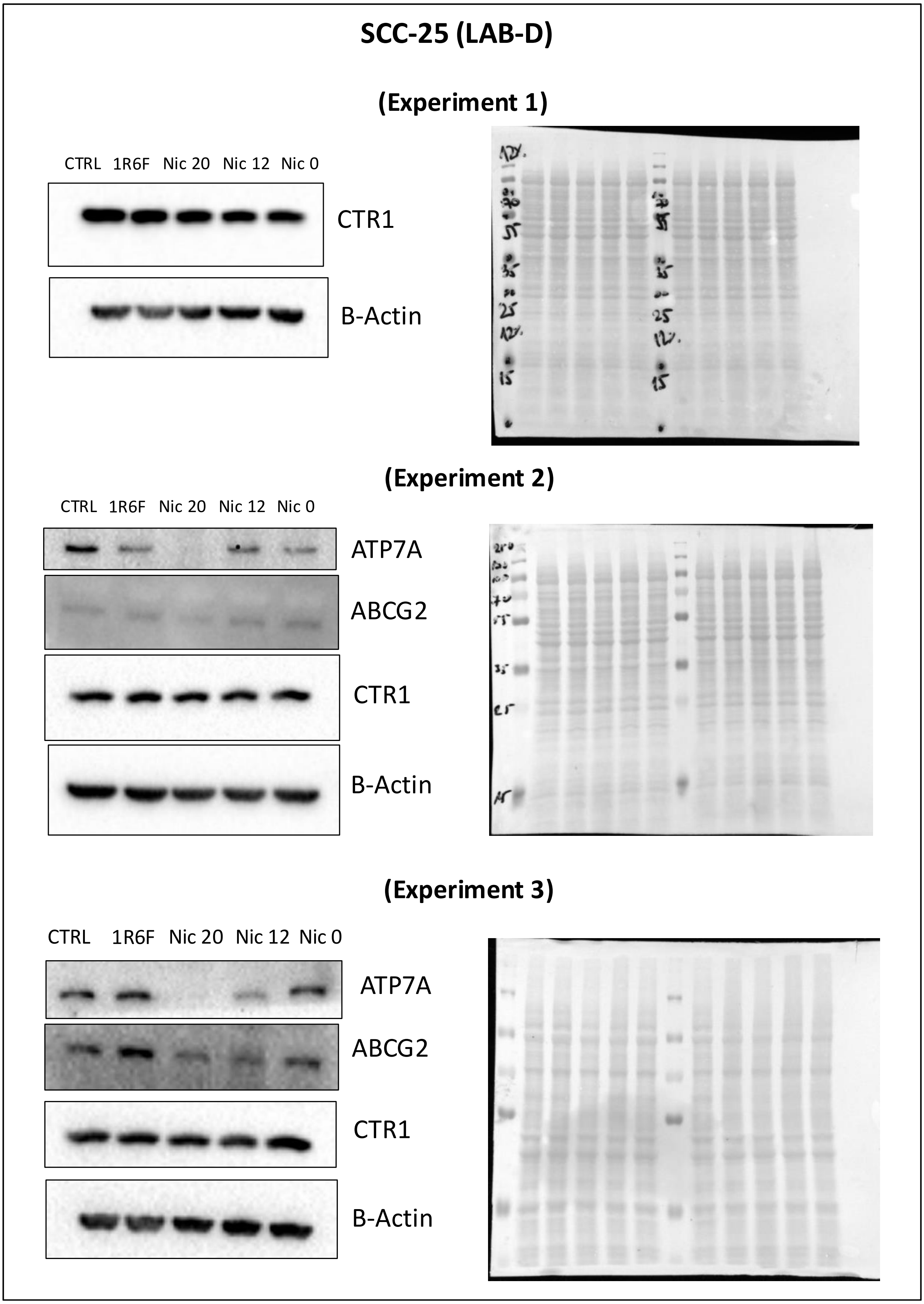


**Figure S2.** Unprocessed blot and the corresponding red ponceau of western blot in SCC-25 cell line executed by LAB-D.

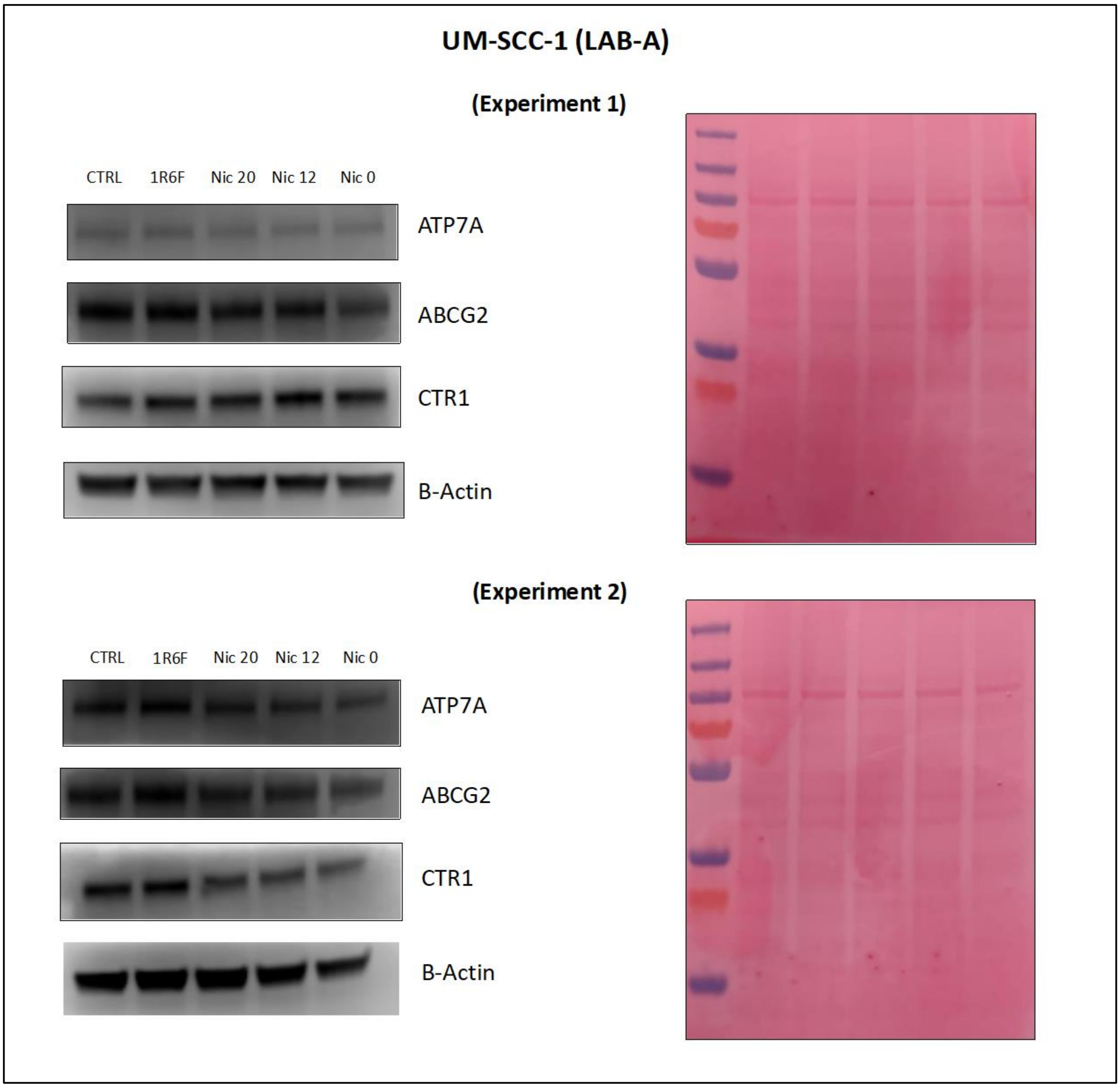


**Figure S3.** Unprocessed blot and the corresponding red ponceau of western blot in UM-SCC-1 cell line executed by LAB-A.

### In cell western blot: plate images

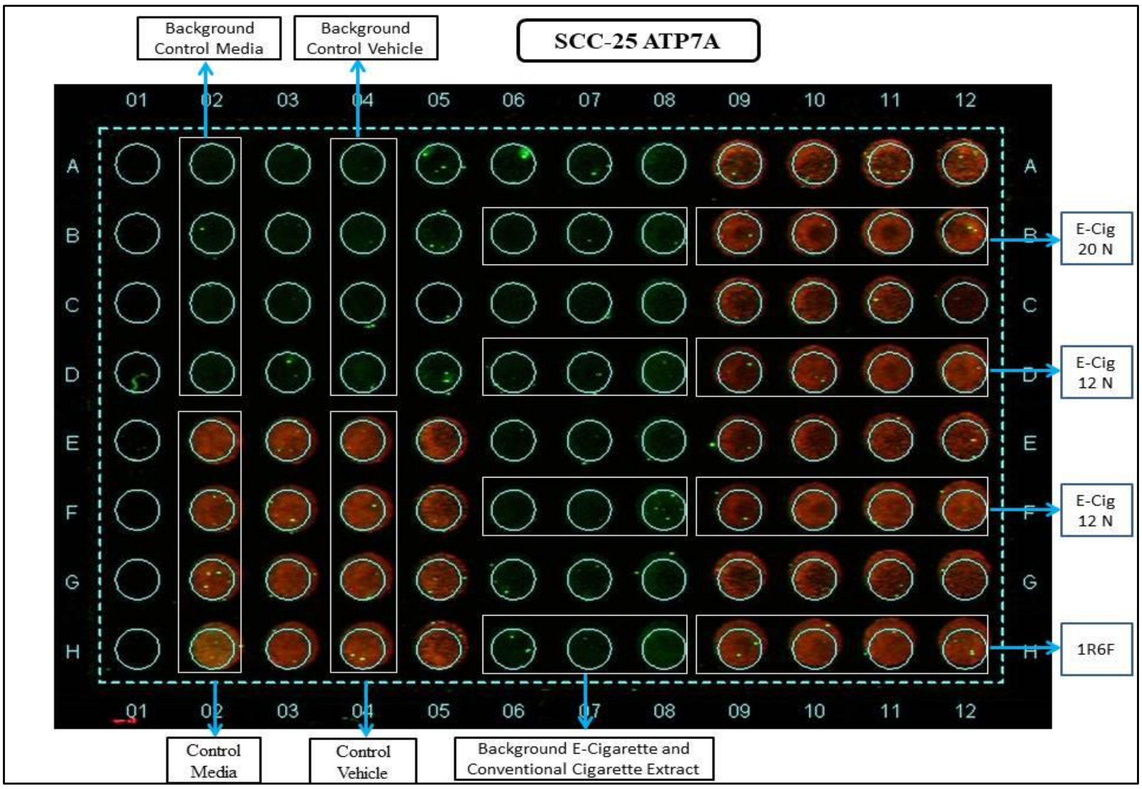


**Figure S4.** Unprocessed image plate of in cell western for ATP7A expression in SCC-25 cell line executed by LAB-C.

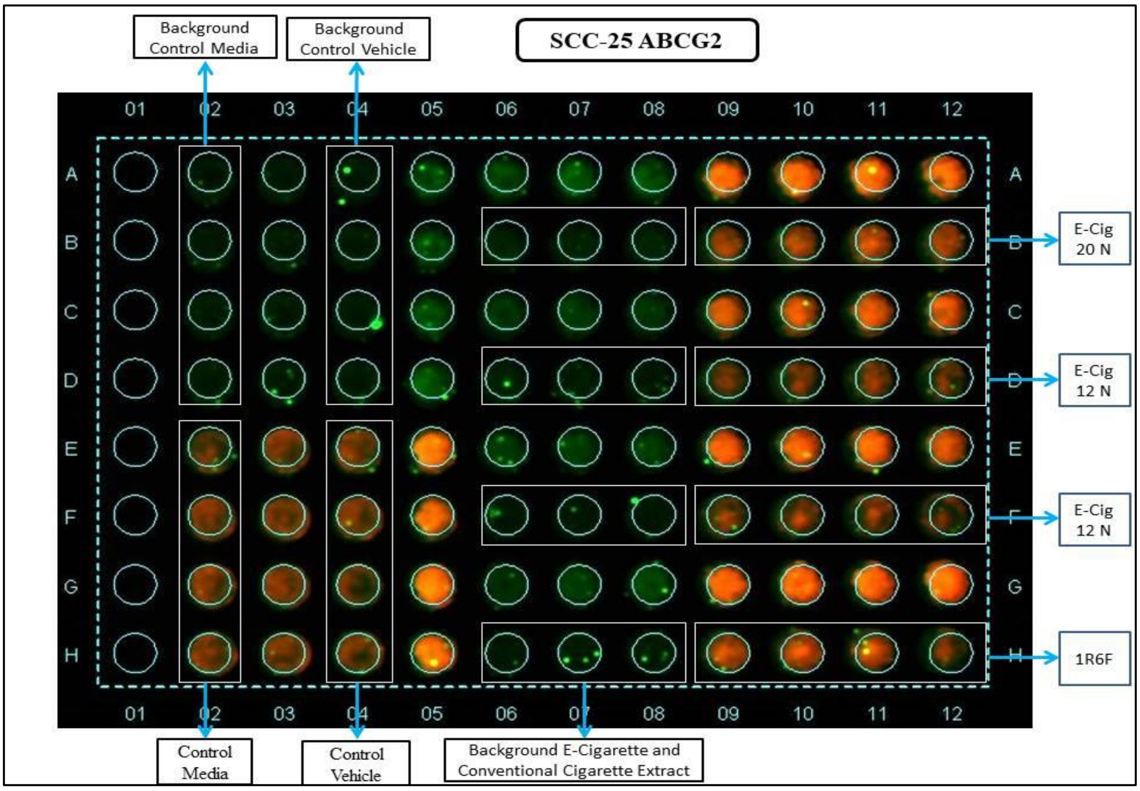


**Figure S5.** Unprocessed image plate of in cell western for ABCG2 expression in SCC-25 cell line executed by LAB-C.

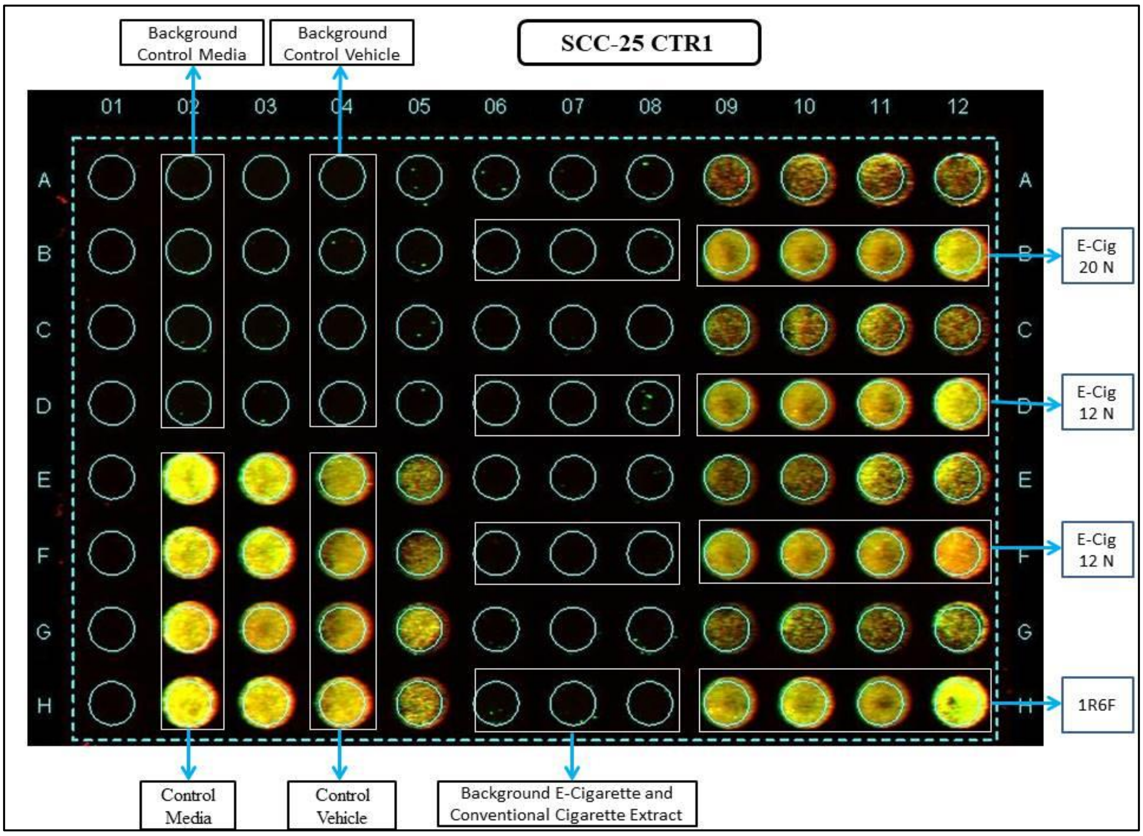


**Figure S6.** Unprocessed image plate of in cell western for CTR1 expression in SCC-25 cell line executed by LAB-C.

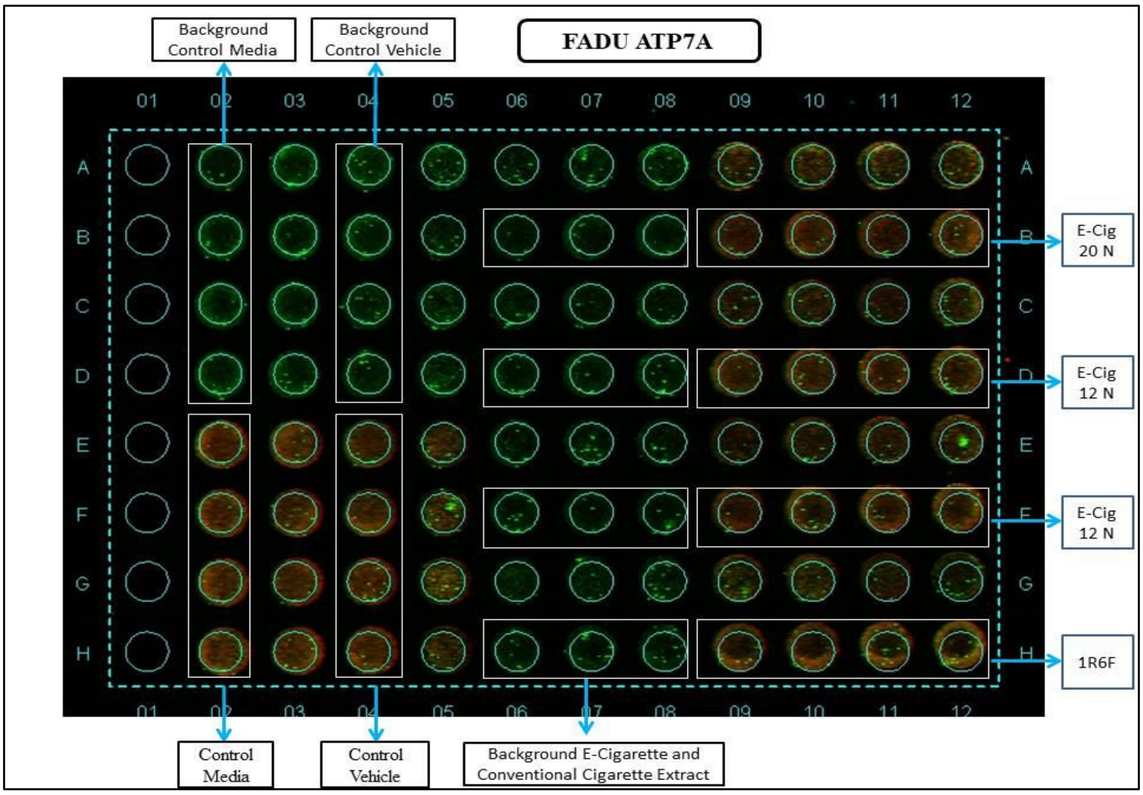


**Figure S7.** Unprocessed image plate of in cell western for ATP7A expression in FaDu cell line executed by LAB-C.

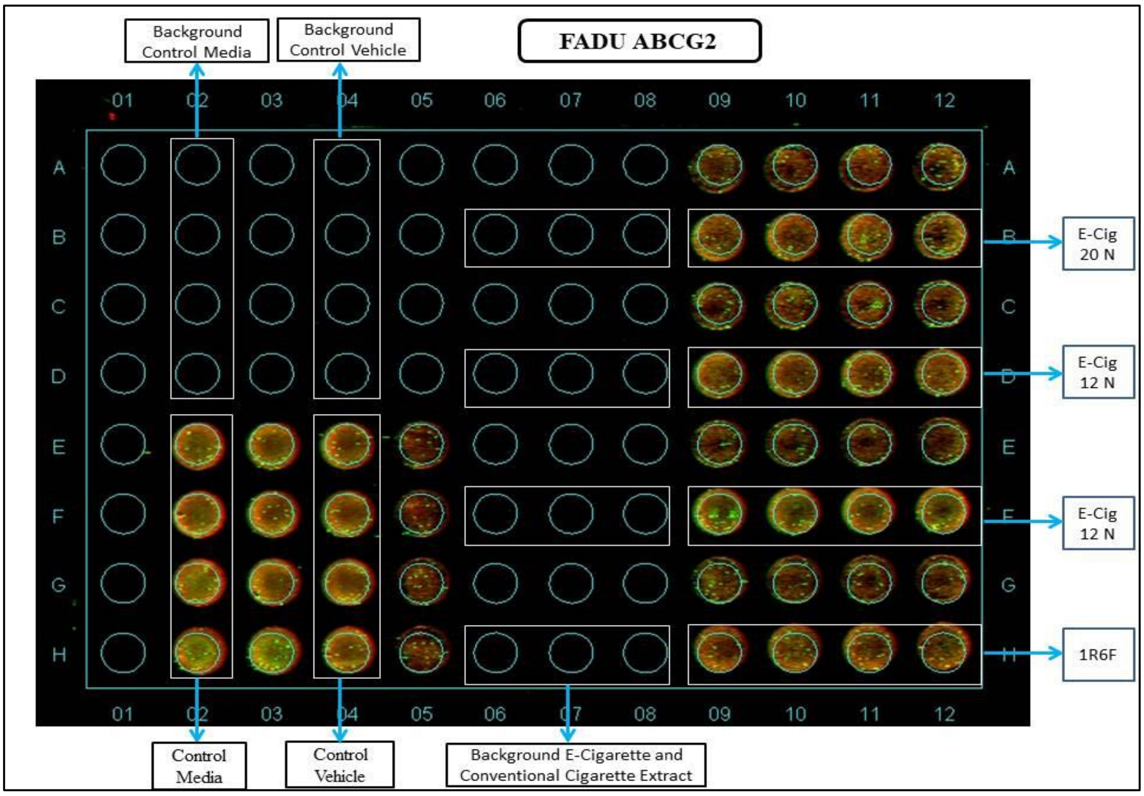


**Figure S8.** Unprocessed image plate of in cell western for ABCG2 expression in FaDu cell line executed by LAB-C.

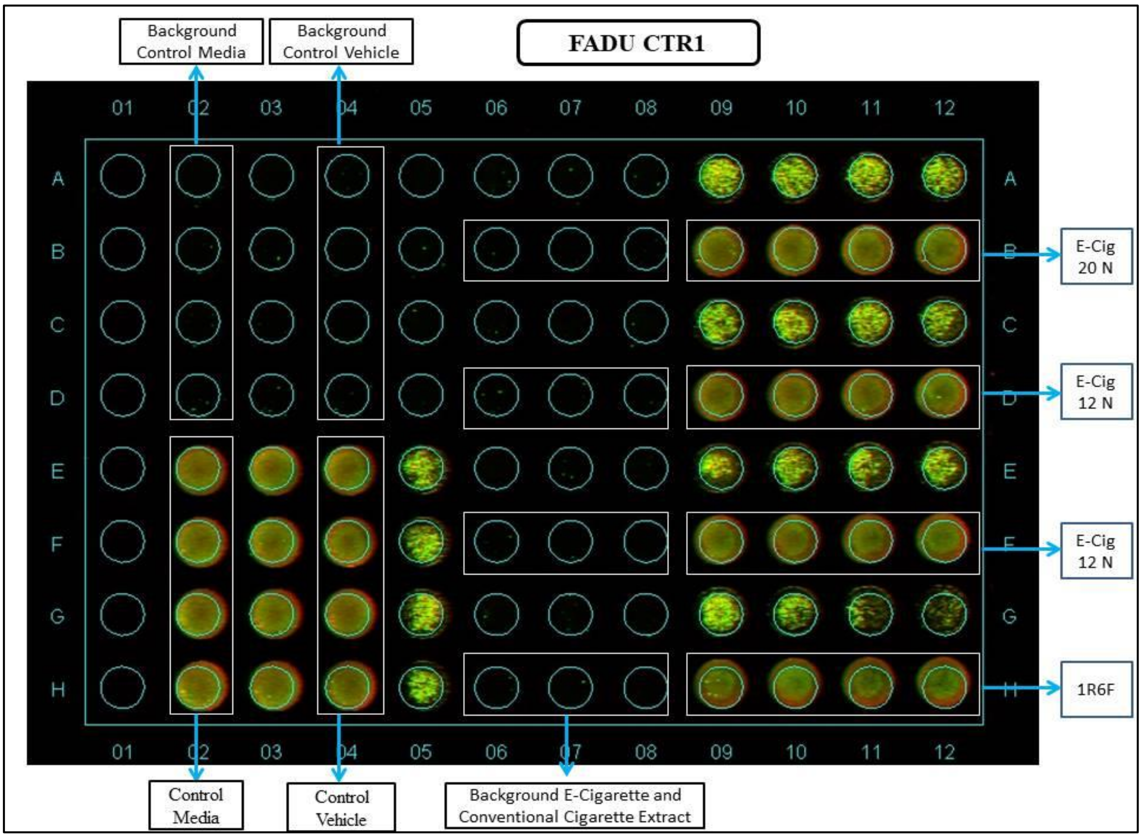


**Figure S9.** Unprocessed image plate of in cell western for CTR1 expression in FaDu cell line executed by LAB-C.

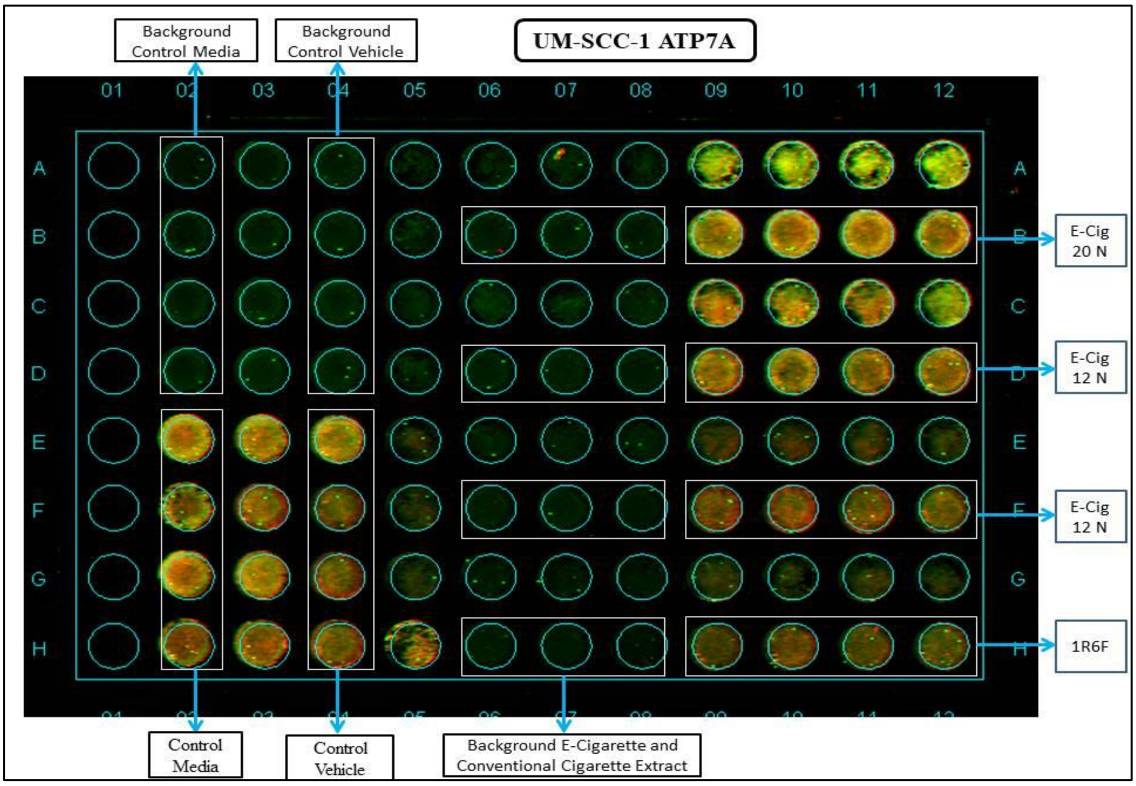


**Figure S10.** Unprocessed image plate of in cell western for ATP7A expression in UM-SCC-1 cell line executed by LAB-C.

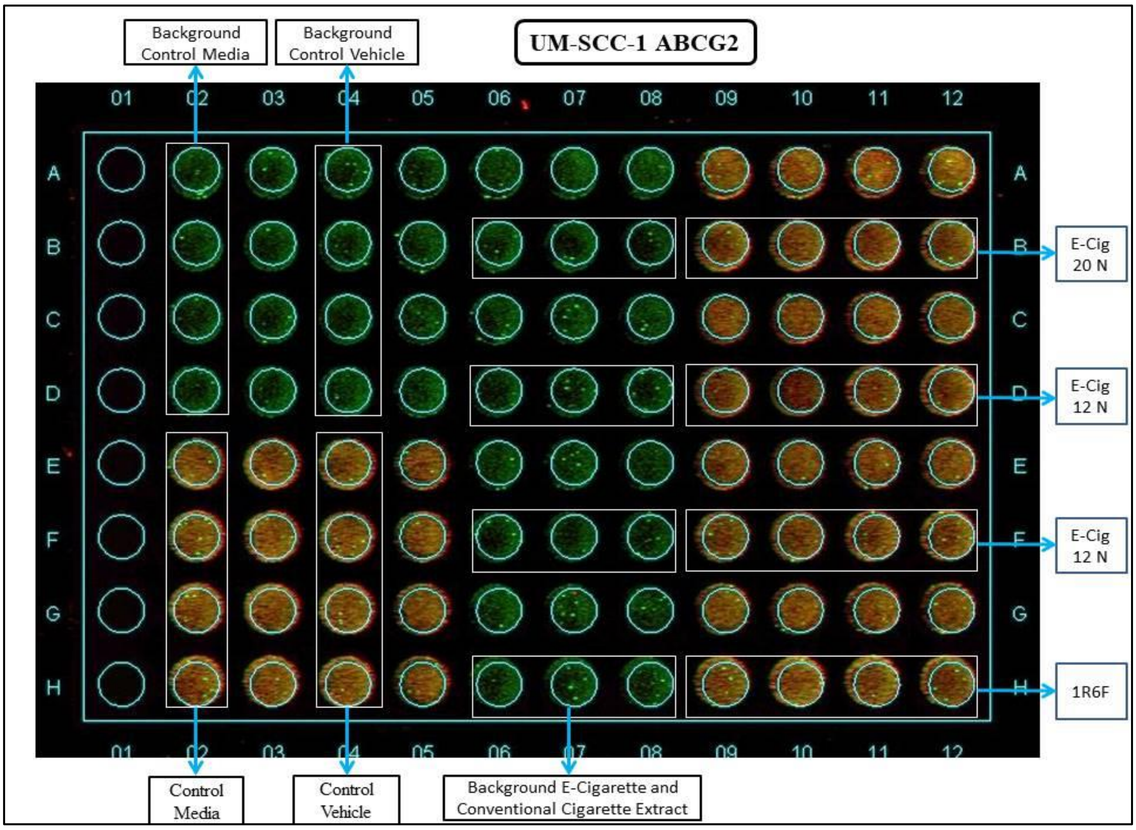


**Figure S11.** Unprocessed image plate of in cell western for ABCG2 expression in UM-SCC-1 cell line executed by LAB-C.

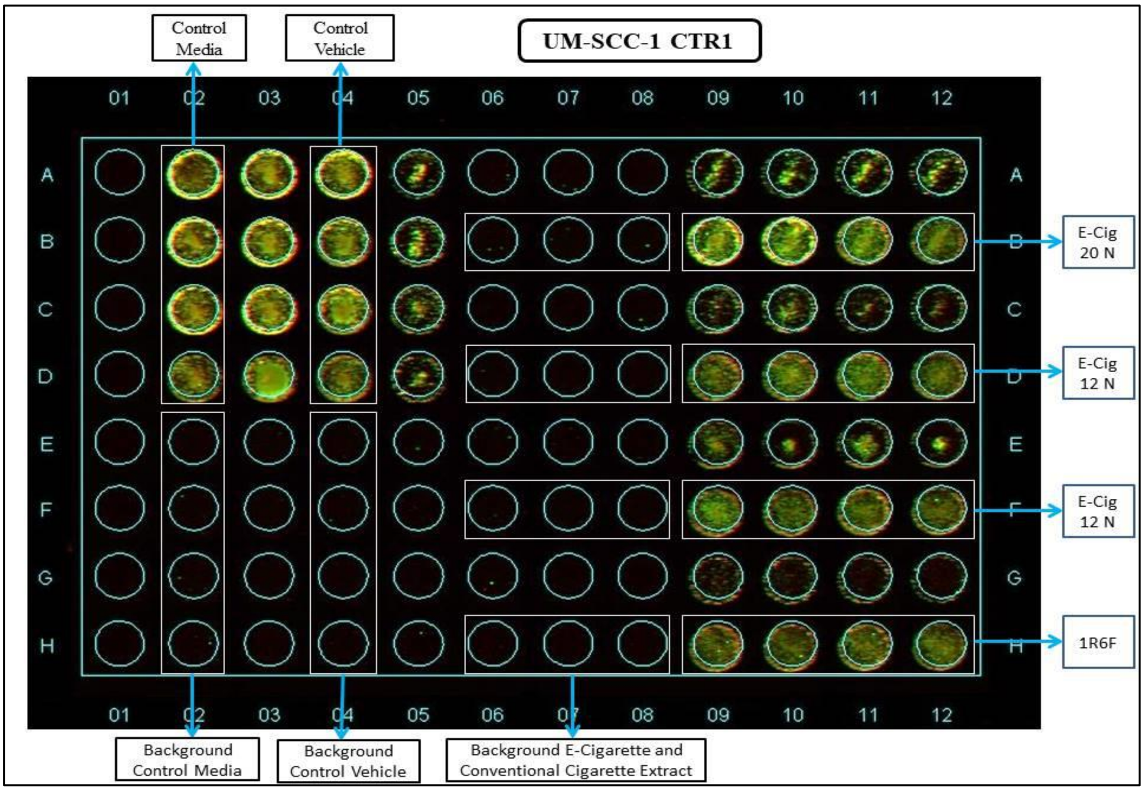


**Figure S12.** Unprocessed image plate of in cell western for CTR1 expression in UM-SCC-1 cell line executed by LAB-C.
